# Digestive system cancer mortality trends by age and sex in Peru, 2001–2020, and projections to 2030

**DOI:** 10.64898/2025.12.17.25342534

**Authors:** Gilmer Solis-Sánchez, Oscar Augusto Lengua-Olivares, Maricela Curisinche-Rojas, Margot Haydee Vidal-Anzardo, Johanny Fidela de Fátima Muro-Cieza, Karina Mayra Aliaga-Llerena de Nuñez

## Abstract

**Background:** Digestive system cancer is a leading cause of mortality worldwide. This study aimed to identify changes in digestive system cancer mortality in Peru (2001–2020) by age and sex, and to predict trends through 2030.

**Methods:** Death records for digestive system cancers (anus, colon, esophagus, stomach, liver and intrahepatic bile ducts, small intestine, pancreas, rectum, biliary tract, and gallbladder) in Peru were analyzed from the World Health Organization mortality database for 2001–2020. Crude mortality rates (CMR) and age-standardized mortality rates (ASMR) per 100,000 inhabitants were calculated by sex, as well as sex ratios and the difference in ASMR between the study period endpoints. Trends were evaluated using Joinpoint regression, and projections to 2030 were performed using the Age-Period-Cohort model with the Nordpred package in R.

**Results:** On average during the period, a digestive system cancer CMR and ASMR of 23.47 and 27.32, respectively, were identified; stomach cancer presented the highest mortality in the study period. During this time, colon cancer surpassed liver cancer as the second leading cause of mortality, with a 65.0% increase in ASMR, representing the largest increase variation between 2001–2020. Conversely, esophageal cancer showed the greatest reduction, with a 25.8% decrease. Regarding projections, we found that between 2021 and 2025, compared to 2020, an increase in ASMR would occur in both sexes only for biliary tract cancer; and this increase is projected to affect six of the ten cancer types during the 2026–2030 period for both sexes.

**Conclusion:** Mortality patterns were heterogeneous, with marked increases observed from 2016 to 2020. Projections suggest a subtle reduction in the 2021–2025 period followed by a resurgence between 2026 and 2030, particularly for colon, stomach, small intestine, pancreas, rectum, and biliary tract cancers. Strengthening prevention and early detection strategies is crucial.

## INTRODUCTION

Cancer is a public health problem worldwide, affecting all countries without exception, although with variations in incidence and mortality rates. These differences depend on the affected tissue, organ, or system. Among the most relevant cancer types are those of the digestive system, which can affect various organs and anatomical zones, including the esophagus, stomach, liver, gallbladder and biliary tract, pancreas, small intestine, colon, rectum, and anus.

In this regard, according to data from the Global Cancer Observatory (Globocan), 4,905,882 new cases of digestive system cancer were registered worldwide in 2022, regardless of sex or age. This corresponds to an age-standardized incidence rate of 47.1 per 100,000 inhabitants, with colon cancer standing out as the most frequent, with 1,142,286 cases and a standardized incidence of 10.7 (1).

Beyond incidence—which undeniably represents a negative impact on public health and healthcare systems—Globocan also reports figures for the Age-Standardized Mortality Rate (ASMR), which amounted to 30.8 per 100,000 inhabitants in 2022, comprising 3,324,774 deaths from digestive system cancer. When evaluated by cancer type, this figure presents a different ranking compared to incidence: liver and bile duct cancer ranks first among deaths (7.4 per 100,000 inhabitants), followed by stomach cancer (6.1 per 100,000) (1).

In Peru, 11,710 deaths from these cancer types have been identified, with an ASMR of 26.3 per 100,000 inhabitants. Specifically, the highest number of deaths corresponds to patients suffering from stomach cancer (n=4,767, ASMR=10.6) (1).

Cancer impacts various aspects of society, one of which encompasses Years of Life Lost (YLL). In 2022, this was determined to be 14.2 years regardless of sex, with a greater magnitude in women (15.3 years) compared to men (13.2 years) (2).

According to the World Health Organization (WHO), cancer mortality is projected to rise to 16.9 million people globally by 2045, representing a significant percentage of deaths in low- and middle-income countries. This is primarily due to a higher prevalence of oncogenic infections, the adoption of Western lifestyles, or inadequate health budget allocation focused on strategic actions other than promotion and prevention; this contrasts with high-income countries, where the increase in mortality is mainly due to higher life expectancy and, consequently, the natural effect of aging as a risk factor for developing cancer. At the regional level, according to the WHO, cancer deaths will increase to 1.35 million in 2045 (80.5% more than in 2022) (3).

Specifically for Peru, the number of cancer deaths is forecast to increase by 16.6% in 2030 compared to 2022, reaching a projection of 13,656 deaths. The cancer with the highest percentage increase in mortality (18.4%) is expected to be gallbladder cancer (from 647 in 2022 to 766 by 2030), followed by liver and bile duct cancer (17.8%), rising from 1,809 deaths in 2022 to 2,131 by 2030 (1).

While specific data on the number of deaths from digestive system cancer over the years is available, it is fundamental to also analyze the ASMR and fluctuations in the trends of these figures in Peru. This will allow for an understanding of the epidemiological dynamics of the disease and generate key information to propose studies that identify its determinants or associated conditions, with the aim of establishing timely and anticipatory control measures (4).

Central and South American countries have experienced significant economic and social changes over the last decades. The reduction in reproduction rates, urbanization, and increased life expectancy are driving major shifts in the demographic structure and associated increases in the burden of non-communicable diseases, including cancer. The cancer profile is changing, and cancers such as prostate and breast cancer are becoming increasingly common in this region, likely reflecting changes in reproductive factors and lifestyles related to economic development (i.e., delayed childbearing, low parity, smoking, alcohol consumption, diets low in fruits and vegetables, obesity, and physical inactivity). As a result, the region faces a “double burden” of cancer, with high rates of infection-related cancers (i.e., cervical, stomach, and liver) and an increase in lifestyle-related (Western) cancers (i.e., prostate, breast, and colorectal).

On the other hand, it should be highlighted that, as previously mentioned, while information exists on predictive mortality estimates performed by Globocan and various studies evaluating this aspect specifically by affected organ or zone (5–7), these studies propose varied methodologies, observation time horizons, and projection lengths. Fundamentally, they do not specifically contemplate data from Peru, representing a significant gap in the body of evidence.

The evaluation of cancer mortality possesses limitations due to the inherent difficulties in registration and access to information, where prospective studies could be designed considering the restrictions inherent to their implementation. However, another strategy—and the one addressed in this research—is to employ the WHO mortality database, which consolidates data reported by different countries regarding numbers, populations, and causes of mortality. Such databases are being used in various investigations pursuing aims and objectives aligned with this research (11–13).

Considering that conditions may exist which modify the epidemiological patterns of pathology distribution, including cancer, and that understanding this can serve to generate predictive estimates useful for proposing health prioritization measures, future planning, and reorientation of resource allocation, the present research sought to identify the trend of the ASMR in Peru by digestive system cancer type during the 2001–2020 period and its projection to the year 2030.

## MATERIALS AND METHODS

### Study design

This study employed an observational, ecological time-series design using secondary data from the World Health Organization (WHO) Mortality Database. This database compiles global death records based on causes of death registered at the national level in compliance with the current International Classification of Diseases (ICD). The WHO Mortality Database collects death counts stratified by country, age group, sex, and year, and is publicly available at: https://www.who.int/data/data-collection-tools/who-mortality-database.

### Study population

The study population comprised death records in Peru for individuals of both sexes and all ages between 2001 and 2020 (the most recent year available at the start of the study). This timeframe was selected as mortality records during this period classified causes of death according to the International Classification of Diseases, 10th Revision (ICD-10). Inclusion was limited to records identifying digestive system neoplasms as the cause of death, corresponding to ICD-10 codes C15–C25; records with unspecified age at the time of death were excluded. Since the study analyzed the entire universe of eligible records, no sample size calculation or sampling procedures were performed.

### Variables

The primary outcome variable was the Age-Standardized Mortality Rate (ASMR), calculated using the direct method based on Segi’s world standard population (14). Age groups were stratified into an initial 0–14 year group, followed by 5-year intervals starting at age 15, expressed per 100,000 person-years.

ASMR was calculated for overall digestive system cancers and stratified by topography: Anus, Colon, Esophagus, Stomach, Liver and intrahepatic bile ducts, Small intestine, Pancreas, Rectum, Biliary tract, and Gallbladder, as detailed in Table 1. All calculations were further stratified by sex.

**Table 1.** Classification of digestive system cancers according to ICD-10.

| Cancer type by topography | ICD-10 codes |
| --- | --- |
| Esophagus | C15 |
| Stomach | C16 |
| Small intestine | C17 |
| Colon | C18-C19 |
| Rectum | C20 |
| Anus | C21 |
| Liver and intrahepatic bile ducts | C22 |
| Gallbladder | C23 |
| Biliary tract | C24 |
| Pancreas | C25 |

**ICD-10:** International Classification of Diseases, 10th Revision.

For the ASMR calculation, the numerator was defined as the number of cancer deaths, and the denominator as the total national population; these calculations were stratified by year, age group, and sex. Population data were obtained from the WHO Mortality Database, sourced from the Population Division of the Department of Economic and Social Affairs of the United Nations Secretariat (https://population.un.org/wpp/). In addition to ASMR, sex, age group, and year of death were included as covariates.

Additionally, the Crude Mortality Rate (CMR) was calculated based solely on death counts and population size, stratified by cancer type and sex.

### Statistical analysis

Since this study utilized publicly available data, no direct primary data collection was performed. Mortality databases were downloaded from the WHO Mortality Database portal (https://www.who.int/data/data-collection-tools/who-mortality-database) and converted to Comma-Separated Values (*.csv) format. Data import, merging, and cleaning were performed using Stata V.17 statistical software (Stata Corporation, College Station, Texas, USA) to generate a specific dataset corresponding to the study population. The processed data were used to calculate ASMRs and death counts, stratified by age group, cancer type, and year of death.

Differences in death counts and ASMRs between the study period endpoints (2001 and 2020) were determined, along with the percentage change calculated as: ([2020 Value - 2001 Value] / 2001 Value) × 100.

Trend evaluation of ASMRs was performed using Joinpoint regression by cancer type using the Joinpoint Desktop Software V.5.3.0.0 (Statistical Research and Applications Branch, National Cancer Institute). The Annual Percent Change (APC) was modeled using log-transformation, assuming constant variance and uncorrelated errors, with 95% empirical quantile confidence intervals (95% CI). Model selection was based on the weighted Bayesian Information Criterion (BIC).

Additionally, ASMR projections to 2030 were performed using RStudio V.12.0, employing an Age-Period-Cohort model with a power-5 link function via the nordpred package. ASMR estimates were generated for five-year periods (2001–2005, 2006–2010, 2011–2015, 2016–2020, 2021–2025, and 2026–2030), accompanied by their respective 95% CIs. A significance level of 0.05 was used for all analyses.

### Ethics

Prior to study execution, the research protocol was evaluated and approved by the Institutional Research Ethics Committee of the National Institute of Health (Instituto Nacional de Salud), under project code OC-019-24. Given that there was no direct contact with human subjects and the data utilized were publicly available in aggregated form without personal identifiers, the requirement for informed consent was waived.

## RESULTS

During the study period, 137,755 deaths were recorded across the different digestive system cancer types. Specifically, the highest number of deaths occurred in stomach cancer cases (n=56,450), while anal cancer had the lowest number of death records (n=567). Furthermore, it was evidenced that from 2003 onwards, the number of annual deaths exceeded 5,000 cases; by 2012, this figure surpassed 7,000, reaching 10,480 in 2020.

During the study period, the average CMR was 23.47, with a slightly higher value in women (24.34) compared to men (22.61); this rate exceeded 30 per 100,000 inhabitants in the last two years of the period. We also found that anal cancer showed the lowest CMR in all years and for the overall period, whereas stomach cancer exhibited the highest values (Table 2).

**Table 2.** CMR by year and digestive system cancer type.

| Digestive system cancer type | CMR by year |  |  |  |  |  |  |  |  |  |  |  |  |  |  |  |  |  |  |  | Overall period |
| --- | --- | --- | --- | --- | --- | --- | --- | --- | --- | --- | --- | --- | --- | --- | --- | --- | --- | --- | --- | --- | --- |
|  | 2001 | 2002 | 2003 | 2004 | 2005 | 2006 | 2007 | 2008 | 2009 | 2010 | 2011 | 2012 | 2013 | 2014 | 2015 | 2016 | 2017 | 2018 | 2019 | 2020 |  |
| Overall - Digestive System |  |  |  |  |  |  |  |  |  |  |  |  |  |  |  |  |  |  |  |  |  |
| Men | 16.13 | 16.46 | 18.16 | 19.83 | 20.98 | 19.67 | 20.51 | 22.62 | 23.03 | 23.57 | 23.55 | 23.05 | 23.19 | 24.05 | 23.70 | 20.83 | 22.97 | 26.13 | 28.83 | 31.39 | 22.61 |
| Women | 17.83 | 18.64 | 19.13 | 21.98 | 24.01 | 20.70 | 23.25 | 24.06 | 25.21 | 25.67 | 24.87 | 24.69 | 24.31 | 25.63 | 24.38 | 21.66 | 25.22 | 27.68 | 31.50 | 32.78 | 24.34 |
| Overall | 16.98 | 17.55 | 18.64 | 20.91 | 22.49 | 20.18 | 21.88 | 23.34 | 24.12 | 24.62 | 24.21 | 23.87 | 23.75 | 24.84 | 24.04 | 21.24 | 24.10 | 26.90 | 30.17 | 32.09 | 23.47 |
| Anus |  |  |  |  |  |  |  |  |  |  |  |  |  |  |  |  |  |  |  |  |  |
| Men | 0.05 | 0.07 | 0.05 | 0.08 | 0.06 | 0.04 | 0.02 | 0.04 | 0.06 | 0.03 | 0.05 | 0.06 | 0.07 | 0.03 | 0.06 | 0.09 | 0.05 | 0.06 | 0.06 | 0.08 | 0.06 |
| Women | 0.10 | 0.11 | 0.07 | 0.13 | 0.10 | 0.11 | 0.08 | 0.08 | 0.10 | 0.12 | 0.13 | 0.12 | 0.15 | 0.22 | 0.17 | 0.14 | 0.18 | 0.15 | 0.15 | 0.27 | 0.14 |
| Overall | 0.08 | 0.09 | 0.06 | 0.10 | 0.08 | 0.08 | 0.05 | 0.06 | 0.08 | 0.07 | 0.09 | 0.09 | 0.11 | 0.13 | 0.12 | 0.11 | 0.12 | 0.11 | 0.11 | 0.17 | 0.10 |
| Colon |  |  |  |  |  |  |  |  |  |  |  |  |  |  |  |  |  |  |  |  |  |
| Men | 1.49 | 1.62 | 1.99 | 2.28 | 2.15 | 2.21 | 2.46 | 2.84 | 2.95 | 3.09 | 3.23 | 3.26 | 3.24 | 3.10 | 3.54 | 2.81 | 3.27 | 3.74 | 4.63 | 5.52 | 3.02 |
| Women | 2.05 | 1.89 | 2.46 | 2.91 | 2.77 | 2.62 | 3.25 | 3.60 | 3.73 | 3.47 | 3.78 | 4.02 | 3.42 | 3.97 | 3.55 | 3.33 | 3.89 | 4.50 | 5.52 | 5.96 | 3.59 |
| Overall | 1.77 | 1.75 | 2.23 | 2.59 | 2.46 | 2.41 | 2.86 | 3.22 | 3.34 | 3.28 | 3.50 | 3.64 | 3.33 | 3.53 | 3.54 | 3.07 | 3.58 | 4.12 | 5.07 | 5.74 | 3.30 |
| Esophagus |  |  |  |  |  |  |  |  |  |  |  |  |  |  |  |  |  |  |  |  |  |
| Men | 0.64 | 0.71 | 0.81 | 0.79 | 0.91 | 1.03 | 0.93 | 0.92 | 0.69 | 1.08 | 0.86 | 0.84 | 0.92 | 0.92 | 0.99 | 0.84 | 0.86 | 1.01 | 0.99 | 0.88 | 0.88 |
| Women | 0.25 | 0.19 | 0.31 | 0.46 | 0.32 | 0.32 | 0.47 | 0.38 | 0.32 | 0.39 | 0.48 | 0.39 | 0.39 | 0.41 | 0.42 | 0.35 | 0.28 | 0.49 | 0.44 | 0.39 | 0.37 |
| Overall | 0.44 | 0.45 | 0.56 | 0.62 | 0.61 | 0.67 | 0.70 | 0.65 | 0.51 | 0.73 | 0.67 | 0.62 | 0.66 | 0.67 | 0.70 | 0.60 | 0.57 | 0.75 | 0.71 | 0.63 | 0.63 |
| Stomach |  |  |  |  |  |  |  |  |  |  |  |  |  |  |  |  |  |  |  |  |  |
| Men | 8.26 | 8.15 | 8.95 | 9.26 | 9.83 | 9.18 | 9.40 | 10.03 | 10.87 | 11.14 | 10.46 | 10.01 | 9.85 | 10.66 | 9.99 | 9.17 | 9.94 | 11.23 | 11.55 | 12.81 | 10.08 |
| Women | 8.26 | 8.30 | 8.35 | 9.08 | 10.24 | 8.00 | 9.12 | 9.15 | 9.63 | 10.36 | 8.90 | 8.56 | 9.09 | 9.32 | 9.14 | 8.00 | 8.69 | 9.28 | 10.11 | 11.09 | 9.15 |
| Overall | 8.26 | 8.23 | 8.65 | 9.17 | 10.03 | 8.59 | 9.26 | 9.59 | 10.25 | 10.75 | 9.68 | 9.28 | 9.47 | 9.99 | 9.56 | 8.58 | 9.31 | 10.25 | 10.83 | 11.94 | 9.62 |
| Liver and intrahepatic bile ducts |  |  |  |  |  |  |  |  |  |  |  |  |  |  |  |  |  |  |  |  |  |
| Men | 2.93 | 3.24 | 3.45 | 3.78 | 4.09 | 3.43 | 3.58 | 4.43 | 4.12 | 3.68 | 4.59 | 4.35 | 4.24 | 4.29 | 4.20 | 3.36 | 3.71 | 4.09 | 4.92 | 4.97 | 3.99 |
| Women | 3.50 | 3.99 | 3.88 | 4.50 | 4.96 | 4.22 | 4.48 | 4.77 | 5.10 | 4.85 | 5.04 | 4.67 | 4.80 | 4.78 | 4.60 | 3.59 | 4.49 | 4.73 | 6.03 | 5.58 | 4.65 |
| Overall | 3.21 | 3.61 | 3.67 | 4.14 | 4.53 | 3.82 | 4.03 | 4.60 | 4.61 | 4.26 | 4.81 | 4.51 | 4.52 | 4.54 | 4.40 | 3.48 | 4.10 | 4.41 | 5.47 | 5.28 | 4.32 |
| Small intestine |  |  |  |  |  |  |  |  |  |  |  |  |  |  |  |  |  |  |  |  |  |
| Men | 0.13 | 0.11 | 0.17 | 0.21 | 0.21 | 0.19 | 0.29 | 0.25 | 0.27 | 0.26 | 0.26 | 0.30 | 0.34 | 0.29 | 0.36 | 0.32 | 0.37 | 0.38 | 0.33 | 0.41 | 0.28 |
| Women | 0.16 | 0.18 | 0.15 | 0.22 | 0.24 | 0.21 | 0.26 | 0.27 | 0.26 | 0.30 | 0.36 | 0.33 | 0.30 | 0.31 | 0.31 | 0.31 | 0.33 | 0.48 | 0.48 | 0.39 | 0.30 |
| Overall | 0.15 | 0.15 | 0.16 | 0.21 | 0.23 | 0.20 | 0.28 | 0.26 | 0.27 | 0.28 | 0.31 | 0.31 | 0.32 | 0.30 | 0.33 | 0.31 | 0.35 | 0.43 | 0.40 | 0.40 | 0.29 |
| Pancreas |  |  |  |  |  |  |  |  |  |  |  |  |  |  |  |  |  |  |  |  |  |
| Men | 1.54 | 1.63 | 1.47 | 1.96 | 1.93 | 1.92 | 2.17 | 2.36 | 2.41 | 2.54 | 2.30 | 2.30 | 2.36 | 2.72 | 2.65 | 2.15 | 2.51 | 3.10 | 3.54 | 3.77 | 2.39 |
| Women | 1.50 | 1.89 | 1.74 | 2.07 | 2.43 | 2.26 | 2.63 | 2.64 | 2.86 | 2.76 | 2.72 | 3.06 | 2.78 | 3.01 | 2.80 | 2.58 | 3.38 | 3.65 | 3.95 | 4.20 | 2.78 |
| Overall | 1.52 | 1.76 | 1.61 | 2.02 | 2.18 | 2.09 | 2.40 | 2.50 | 2.63 | 2.65 | 2.51 | 2.68 | 2.57 | 2.87 | 2.73 | 2.36 | 2.95 | 3.38 | 3.75 | 3.98 | 2.59 |
| Rectum |  |  |  |  |  |  |  |  |  |  |  |  |  |  |  |  |  |  |  |  |  |
| Men | 0.32 | 0.32 | 0.41 | 0.37 | 0.62 | 0.50 | 0.51 | 0.46 | 0.46 | 0.59 | 0.50 | 0.54 | 0.68 | 0.62 | 0.68 | 0.65 | 0.61 | 0.69 | 0.83 | 0.94 | 0.57 |
| Women | 0.30 | 0.30 | 0.30 | 0.43 | 0.47 | 0.44 | 0.46 | 0.57 | 0.48 | 0.68 | 0.58 | 0.69 | 0.59 | 0.64 | 0.65 | 0.57 | 0.60 | 0.79 | 0.70 | 0.88 | 0.56 |
| Overall | 0.31 | 0.31 | 0.35 | 0.40 | 0.54 | 0.47 | 0.48 | 0.51 | 0.47 | 0.64 | 0.54 | 0.62 | 0.64 | 0.63 | 0.66 | 0.61 | 0.61 | 0.74 | 0.77 | 0.91 | 0.57 |
| Biliary tract |  |  |  |  |  |  |  |  |  |  |  |  |  |  |  |  |  |  |  |  |  |
|  | 2001 | 2002 | 2003 | 2004 | 2005 | 2006 | 2007 | 2008 | 2009 | 2010 | 2011 | 2012 | 2013 | 2014 | 2015 | 2016 | 2017 | 2018 | 2019 | 2020 |  |
| Men | 0.38 | 0.24 | 0.41 | 0.64 | 0.63 | 0.58 | 0.62 | 0.59 | 0.58 | 0.59 | 0.64 | 0.74 | 0.85 | 0.79 | 0.63 | 0.90 | 1.11 | 1.04 | 1.29 | 1.23 | 0.74 |
| Women | 0.61 | 0.62 | 0.66 | 0.99 | 1.04 | 1.07 | 0.97 | 1.02 | 1.11 | 1.13 | 1.16 | 1.22 | 1.19 | 1.25 | 1.09 | 1.28 | 1.61 | 1.55 | 1.87 | 1.76 | 1.18 |
| Overall | 0.50 | 0.43 | 0.53 | 0.82 | 0.83 | 0.82 | 0.79 | 0.80 | 0.85 | 0.86 | 0.90 | 0.98 | 1.02 | 1.02 | 0.86 | 1.09 | 1.36 | 1.30 | 1.59 | 1.50 | 0.96 |
| Gallbladder |  |  |  |  |  |  |  |  |  |  |  |  |  |  |  |  |  |  |  |  |  |
| Men | 0.38 | 0.38 | 0.45 | 0.46 | 0.55 | 0.59 | 0.53 | 0.70 | 0.62 | 0.57 | 0.67 | 0.65 | 0.63 | 0.62 | 0.62 | 0.54 | 0.54 | 0.78 | 0.68 | 0.79 | 0.59 |
| Women | 1.10 | 1.17 | 1.20 | 1.20 | 1.45 | 1.45 | 1.53 | 1.59 | 1.63 | 1.60 | 1.72 | 1.62 | 1.60 | 1.73 | 1.64 | 1.50 | 1.76 | 2.05 | 2.24 | 2.26 | 1.62 |
| Overall | 0.74 | 0.77 | 0.82 | 0.83 | 1.00 | 1.02 | 1.03 | 1.15 | 1.12 | 1.09 | 1.20 | 1.14 | 1.12 | 1.18 | 1.13 | 1.02 | 1.15 | 1.42 | 1.46 | 1.53 | 1.11 |
| CMR: Crude mortality rate. |  |  |  |  |  |  |  |  |  |  |  |  |  |  |  |  |  |  |  |  |  |

In the evaluation of ASMR, an overall average of 27.32 was identified for digestive system cancers. This measure presented an irregular pattern of reduction until 2016, followed by an increase up to 2020. The highest ASMR by digestive cancer type was observed for stomach cancer, with a value of 11.37, followed by liver and intrahepatic bile duct cancer with an ASMR of 5.06 (Table 3).

**Table 3.**
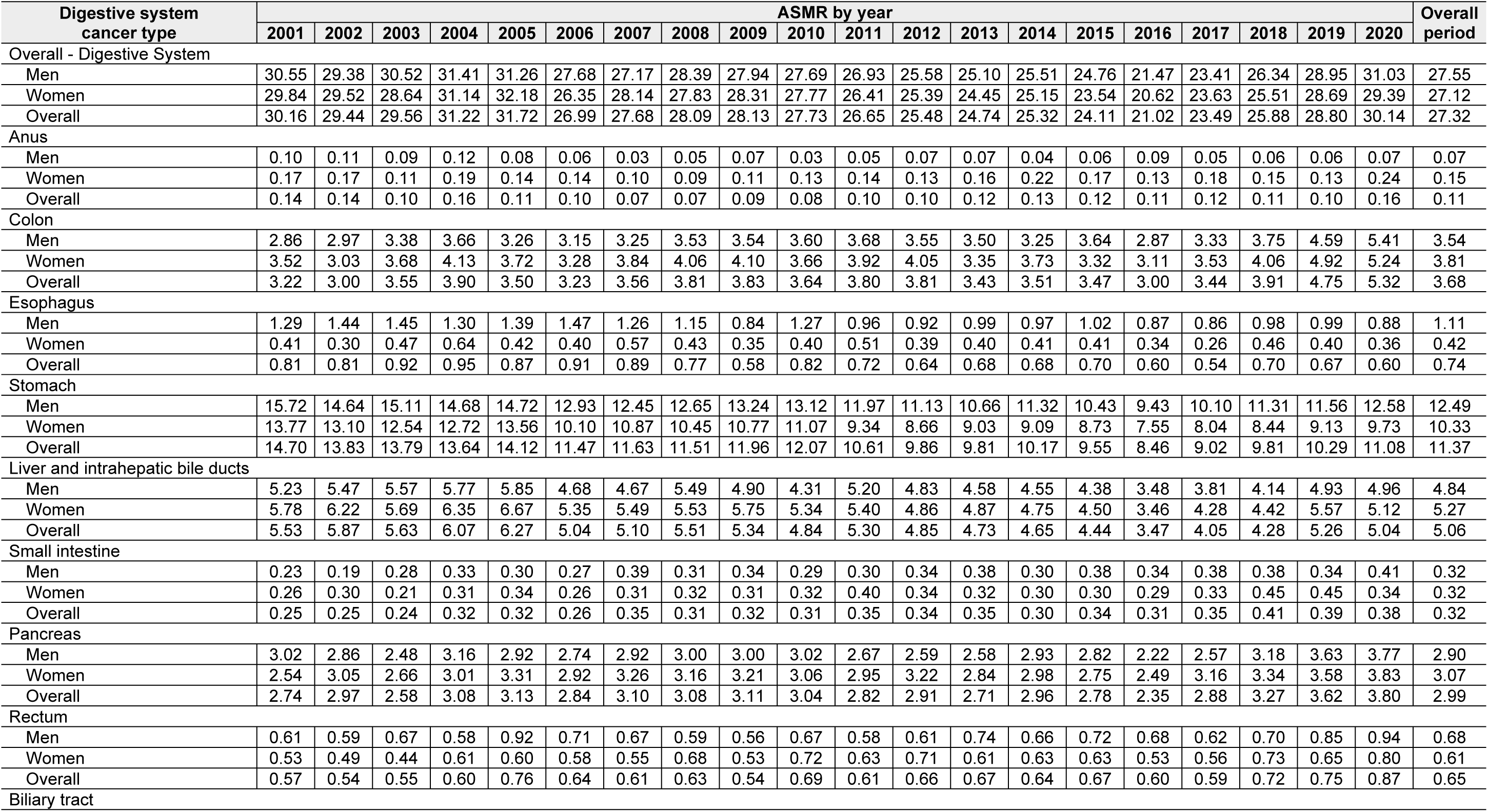
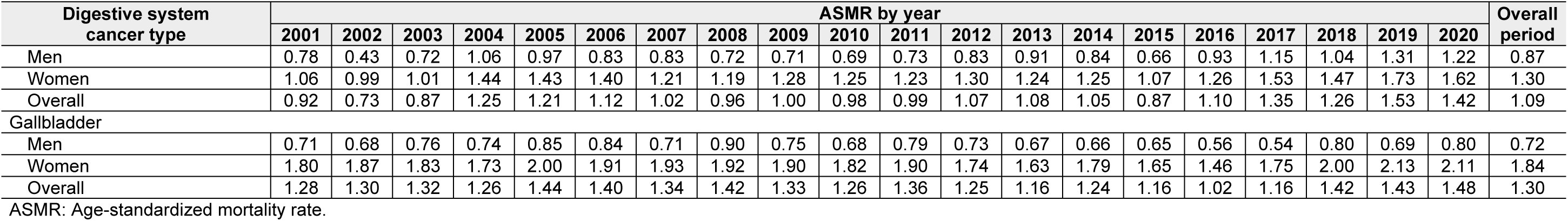
ASMR by year and digestive system cancer type.

| Digestive system cancer type | ASMR by year |  |  |  |  |  |  |  |  |  |  |  |  |  |  |  |  |  |  |  | Overall period |
| --- | --- | --- | --- | --- | --- | --- | --- | --- | --- | --- | --- | --- | --- | --- | --- | --- | --- | --- | --- | --- | --- |
|  | 2001 | 2002 | 2003 | 2004 | 2005 | 2006 | 2007 | 2008 | 2009 | 2010 | 2011 | 2012 | 2013 | 2014 | 2015 | 2016 | 2017 | 2018 | 2019 | 2020 |  |
| Overall - Digestive System |  |  |  |  |  |  |  |  |  |  |  |  |  |  |  |  |  |  |  |  |  |
| Men | 30.55 | 29.38 | 30.52 | 31.41 | 31.26 | 27.68 | 27.17 | 28.39 | 27.94 | 27.69 | 26.93 | 25.58 | 25.10 | 25.51 | 24.76 | 21.47 | 23.41 | 26.34 | 28.95 | 31.03 | 27.55 |
| Women | 29.84 | 29.52 | 28.64 | 31.14 | 32.18 | 26.35 | 28.14 | 27.83 | 28.31 | 27.77 | 26.41 | 25.39 | 24.45 | 25.15 | 23.54 | 20.62 | 23.63 | 25.51 | 28.69 | 29.39 | 27.12 |
| Overall | 30.16 | 29.44 | 29.56 | 31.22 | 31.72 | 26.99 | 27.68 | 28.09 | 28.13 | 27.73 | 26.65 | 25.48 | 24.74 | 25.32 | 24.11 | 21.02 | 23.49 | 25.88 | 28.80 | 30.14 | 27.32 |
| Anus |  |  |  |  |  |  |  |  |  |  |  |  |  |  |  |  |  |  |  |  |  |
| Men | 0.10 | 0.11 | 0.09 | 0.12 | 0.08 | 0.06 | 0.03 | 0.05 | 0.07 | 0.03 | 0.05 | 0.07 | 0.07 | 0.04 | 0.06 | 0.09 | 0.05 | 0.06 | 0.06 | 0.07 | 0.07 |
| Women | 0.17 | 0.17 | 0.11 | 0.19 | 0.14 | 0.14 | 0.10 | 0.09 | 0.11 | 0.13 | 0.14 | 0.13 | 0.16 | 0.22 | 0.17 | 0.13 | 0.18 | 0.15 | 0.13 | 0.24 | 0.15 |
| Overall | 0.14 | 0.14 | 0.10 | 0.16 | 0.11 | 0.10 | 0.07 | 0.07 | 0.09 | 0.08 | 0.10 | 0.10 | 0.12 | 0.13 | 0.12 | 0.11 | 0.12 | 0.11 | 0.10 | 0.16 | 0.11 |
| Colon |  |  |  |  |  |  |  |  |  |  |  |  |  |  |  |  |  |  |  |  |  |
| Men | 2.86 | 2.97 | 3.38 | 3.66 | 3.26 | 3.15 | 3.25 | 3.53 | 3.54 | 3.60 | 3.68 | 3.55 | 3.50 | 3.25 | 3.64 | 2.87 | 3.33 | 3.75 | 4.59 | 5.41 | 3.54 |
| Women | 3.52 | 3.03 | 3.68 | 4.13 | 3.72 | 3.28 | 3.84 | 4.06 | 4.10 | 3.66 | 3.92 | 4.05 | 3.35 | 3.73 | 3.32 | 3.11 | 3.53 | 4.06 | 4.92 | 5.24 | 3.81 |
| Overall | 3.22 | 3.00 | 3.55 | 3.90 | 3.50 | 3.23 | 3.56 | 3.81 | 3.83 | 3.64 | 3.80 | 3.81 | 3.43 | 3.51 | 3.47 | 3.00 | 3.44 | 3.91 | 4.75 | 5.32 | 3.68 |
| Esophagus |  |  |  |  |  |  |  |  |  |  |  |  |  |  |  |  |  |  |  |  |  |
| Men | 1.29 | 1.44 | 1.45 | 1.30 | 1.39 | 1.47 | 1.26 | 1.15 | 0.84 | 1.27 | 0.96 | 0.92 | 0.99 | 0.97 | 1.02 | 0.87 | 0.86 | 0.98 | 0.99 | 0.88 | 1.11 |
| Women | 0.41 | 0.30 | 0.47 | 0.64 | 0.42 | 0.40 | 0.57 | 0.43 | 0.35 | 0.40 | 0.51 | 0.39 | 0.40 | 0.41 | 0.41 | 0.34 | 0.26 | 0.46 | 0.40 | 0.36 | 0.42 |
| Overall | 0.81 | 0.81 | 0.92 | 0.95 | 0.87 | 0.91 | 0.89 | 0.77 | 0.58 | 0.82 | 0.72 | 0.64 | 0.68 | 0.68 | 0.70 | 0.60 | 0.54 | 0.70 | 0.67 | 0.60 | 0.74 |
| Stomach |  |  |  |  |  |  |  |  |  |  |  |  |  |  |  |  |  |  |  |  |  |
| Men | 15.72 | 14.64 | 15.11 | 14.68 | 14.72 | 12.93 | 12.45 | 12.65 | 13.24 | 13.12 | 11.97 | 11.13 | 10.66 | 11.32 | 10.43 | 9.43 | 10.10 | 11.31 | 11.56 | 12.58 | 12.49 |
| Women | 13.77 | 13.10 | 12.54 | 12.72 | 13.56 | 10.10 | 10.87 | 10.45 | 10.77 | 11.07 | 9.34 | 8.66 | 9.03 | 9.09 | 8.73 | 7.55 | 8.04 | 8.44 | 9.13 | 9.73 | 10.33 |
| Overall | 14.70 | 13.83 | 13.79 | 13.64 | 14.12 | 11.47 | 11.63 | 11.51 | 11.96 | 12.07 | 10.61 | 9.86 | 9.81 | 10.17 | 9.55 | 8.46 | 9.02 | 9.81 | 10.29 | 11.08 | 11.37 |
| Liver and intrahepatic bile ducts |  |  |  |  |  |  |  |  |  |  |  |  |  |  |  |  |  |  |  |  |  |
| Men | 5.23 | 5.47 | 5.57 | 5.77 | 5.85 | 4.68 | 4.67 | 5.49 | 4.90 | 4.31 | 5.20 | 4.83 | 4.58 | 4.55 | 4.38 | 3.48 | 3.81 | 4.14 | 4.93 | 4.96 | 4.84 |
| Women | 5.78 | 6.22 | 5.69 | 6.35 | 6.67 | 5.35 | 5.49 | 5.53 | 5.75 | 5.34 | 5.40 | 4.86 | 4.87 | 4.75 | 4.50 | 3.46 | 4.28 | 4.42 | 5.57 | 5.12 | 5.27 |
| Overall | 5.53 | 5.87 | 5.63 | 6.07 | 6.27 | 5.04 | 5.10 | 5.51 | 5.34 | 4.84 | 5.30 | 4.85 | 4.73 | 4.65 | 4.44 | 3.47 | 4.05 | 4.28 | 5.26 | 5.04 | 5.06 |
| Small intestine |  |  |  |  |  |  |  |  |  |  |  |  |  |  |  |  |  |  |  |  |  |
| Men | 0.23 | 0.19 | 0.28 | 0.33 | 0.30 | 0.27 | 0.39 | 0.31 | 0.34 | 0.29 | 0.30 | 0.34 | 0.38 | 0.30 | 0.38 | 0.34 | 0.38 | 0.38 | 0.34 | 0.41 | 0.32 |
| Women | 0.26 | 0.30 | 0.21 | 0.31 | 0.34 | 0.26 | 0.31 | 0.32 | 0.31 | 0.32 | 0.40 | 0.34 | 0.32 | 0.30 | 0.30 | 0.29 | 0.33 | 0.45 | 0.45 | 0.34 | 0.32 |
| Overall | 0.25 | 0.25 | 0.24 | 0.32 | 0.32 | 0.26 | 0.35 | 0.31 | 0.32 | 0.31 | 0.35 | 0.34 | 0.35 | 0.30 | 0.34 | 0.31 | 0.35 | 0.41 | 0.39 | 0.38 | 0.32 |
| Pancreas |  |  |  |  |  |  |  |  |  |  |  |  |  |  |  |  |  |  |  |  |  |
| Men | 3.02 | 2.86 | 2.48 | 3.16 | 2.92 | 2.74 | 2.92 | 3.00 | 3.00 | 3.02 | 2.67 | 2.59 | 2.58 | 2.93 | 2.82 | 2.22 | 2.57 | 3.18 | 3.63 | 3.77 | 2.90 |
| Women | 2.54 | 3.05 | 2.66 | 3.01 | 3.31 | 2.92 | 3.26 | 3.16 | 3.21 | 3.06 | 2.95 | 3.22 | 2.84 | 2.98 | 2.75 | 2.49 | 3.16 | 3.34 | 3.58 | 3.83 | 3.07 |
| Overall | 2.74 | 2.97 | 2.58 | 3.08 | 3.13 | 2.84 | 3.10 | 3.08 | 3.11 | 3.04 | 2.82 | 2.91 | 2.71 | 2.96 | 2.78 | 2.35 | 2.88 | 3.27 | 3.62 | 3.80 | 2.99 |
| Rectum |  |  |  |  |  |  |  |  |  |  |  |  |  |  |  |  |  |  |  |  |  |
| Men | 0.61 | 0.59 | 0.67 | 0.58 | 0.92 | 0.71 | 0.67 | 0.59 | 0.56 | 0.67 | 0.58 | 0.61 | 0.74 | 0.66 | 0.72 | 0.68 | 0.62 | 0.70 | 0.85 | 0.94 | 0.68 |
| Women | 0.53 | 0.49 | 0.44 | 0.61 | 0.60 | 0.58 | 0.55 | 0.68 | 0.53 | 0.72 | 0.63 | 0.71 | 0.61 | 0.63 | 0.63 | 0.53 | 0.56 | 0.73 | 0.65 | 0.80 | 0.61 |
| Overall | 0.57 | 0.54 | 0.55 | 0.60 | 0.76 | 0.64 | 0.61 | 0.63 | 0.54 | 0.69 | 0.61 | 0.66 | 0.67 | 0.64 | 0.67 | 0.60 | 0.59 | 0.72 | 0.75 | 0.87 | 0.65 |
| Biliary tract |  |  |  |  |  |  |  |  |  |  |  |  |  |  |  |  |  |  |  |  |  |

| Digestive system<br>cancer type | ASMR by year |  |  |  |  |  |  |  |  |  |  |  |  |  |  |  |  |  |  |  | Overall<br>period |
| --- | --- | --- | --- | --- | --- | --- | --- | --- | --- | --- | --- | --- | --- | --- | --- | --- | --- | --- | --- | --- | --- |
|  | 2001 | 2002 | 2003 | 2004 | 2005 | 2006 | 2007 | 2008 | 2009 | 2010 | 2011 | 2012 | 2013 | 2014 | 2015 | 2016 | 2017 | 2018 | 2019 | 2020 |  |
| Men | 0.78 | 0.43 | 0.72 | 1.06 | 0.97 | 0.83 | 0.83 | 0.72 | 0.71 | 0.69 | 0.73 | 0.83 | 0.91 | 0.84 | 0.66 | 0.93 | 1.15 | 1.04 | 1.31 | 1.22 | 0.87 |
| Women | 1.06 | 0.99 | 1.01 | 1.44 | 1.43 | 1.40 | 1.21 | 1.19 | 1.28 | 1.25 | 1.23 | 1.30 | 1.24 | 1.25 | 1.07 | 1.26 | 1.53 | 1.47 | 1.73 | 1.62 | 1.30 |
| Overall | 0.92 | 0.73 | 0.87 | 1.25 | 1.21 | 1.12 | 1.02 | 0.96 | 1.00 | 0.98 | 0.99 | 1.07 | 1.08 | 1.05 | 0.87 | 1.10 | 1.35 | 1.26 | 1.53 | 1.42 | 1.09 |
| Gallbladder |  |  |  |  |  |  |  |  |  |  |  |  |  |  |  |  |  |  |  |  |  |
| Men | 0.71 | 0.68 | 0.76 | 0.74 | 0.85 | 0.84 | 0.71 | 0.90 | 0.75 | 0.68 | 0.79 | 0.73 | 0.67 | 0.66 | 0.65 | 0.56 | 0.54 | 0.80 | 0.69 | 0.80 | 0.72 |
| Women | 1.80 | 1.87 | 1.83 | 1.73 | 2.00 | 1.91 | 1.93 | 1.92 | 1.90 | 1.82 | 1.90 | 1.74 | 1.63 | 1.79 | 1.65 | 1.46 | 1.75 | 2.00 | 2.13 | 2.11 | 1.84 |
| Overall | 1.28 | 1.30 | 1.32 | 1.26 | 1.44 | 1.40 | 1.34 | 1.42 | 1.33 | 1.26 | 1.36 | 1.25 | 1.16 | 1.24 | 1.16 | 1.02 | 1.16 | 1.42 | 1.43 | 1.48 | 1.30 |

ASMR: Age-standardized mortality rate.

Assessing ASMR patterns by sex, distinct presentations were identified for anal, esophageal, biliary tract, and gallbladder cancers. Specifically, the male-to-female ASMR ratio for gallbladder cancer was 0.39, evidencing higher mortality in women; a similar pattern was observed for anal cancer, with an ASMR ratio of 0.46. In contrast, esophageal cancer mortality was higher in men, with an ASMR ratio of 2.67. The overall period ratio for digestive system cancers was 1.02 (Table 4).

**Table 4.** ASMR sex ratio by year and digestive system cancer type.

| Digestive system cancer type | ASMR sex ratio |  |  |  |  |  |  |  |  |  |  |  |  |  |  |  |  |  |  |  | Overall<br>I<br>period |
| --- | --- | --- | --- | --- | --- | --- | --- | --- | --- | --- | --- | --- | --- | --- | --- | --- | --- | --- | --- | --- | --- |
|  | 200<br>1 | 200<br>2 | 200<br>3 | 200<br>4 | 200<br>5 | 200<br>6 | 200<br>7 | 200<br>8 | 200<br>9 | 201<br>0 | 201<br>1 | 201<br>2 | 201<br>3 | 201<br>4 | 201<br>5 | 201<br>6 | 201<br>7 | 201<br>8 | 201<br>9 | 202<br>0 |  |
| Overall - Digestive System | 1.02 | 1.00 | 1.07 | 1.01 | 0.97 | 1.05 | 0.97 | 1.02 | 0.99 | 1.00 | 1.02 | 1.01 | 1.03 | 1.01 | 1.05 | 1.04 | 0.99 | 1.03 | 1.01 | 1.06 | 1.02 |
| Anus | 0.61 | 0.65 | 0.82 | 0.64 | 0.61 | 0.41 | 0.26 | 0.54 | 0.61 | 0.24 | 0.39 | 0.56 | 0.47 | 0.17 | 0.38 | 0.67 | 0.30 | 0.41 | 0.48 | 0.30 | 0.46 |
| Colon | 0.81 | 0.98 | 0.92 | 0.89 | 0.88 | 0.96 | 0.85 | 0.87 | 0.86 | 0.98 | 0.94 | 0.88 | 1.05 | 0.87 | 1.10 | 0.92 | 0.94 | 0.92 | 0.93 | 1.03 | 0.93 |
| Esophagus | 3.12 | 4.73 | 3.07 | 2.03 | 3.31 | 3.65 | 2.22 | 2.64 | 2.37 | 3.19 | 1.90 | 2.39 | 2.46 | 2.34 | 2.46 | 2.58 | 3.24 | 2.14 | 2.49 | 2.45 | 2.67 |
| Stomach | 1.14 | 1.12 | 1.21 | 1.15 | 1.09 | 1.28 | 1.15 | 1.21 | 1.23 | 1.19 | 1.28 | 1.28 | 1.18 | 1.25 | 1.19 | 1.25 | 1.26 | 1.34 | 1.27 | 1.29 | 1.21 |
| Liver and intrahepatic bile ducts | 0.90 | 0.88 | 0.98 | 0.91 | 0.88 | 0.88 | 0.85 | 0.99 | 0.85 | 0.81 | 0.96 | 0.99 | 0.94 | 0.96 | 0.97 | 1.01 | 0.89 | 0.94 | 0.88 | 0.97 | 0.92 |
| Small intestine | 0.90 | 0.64 | 1.34 | 1.06 | 0.86 | 1.04 | 1.25 | 0.97 | 1.10 | 0.91 | 0.75 | 0.98 | 1.19 | 0.98 | 1.25 | 1.16 | 1.18 | 0.84 | 0.75 | 1.20 | 1.00 |
| Pancreas | 1.19 | 0.94 | 0.93 | 1.05 | 0.88 | 0.94 | 0.90 | 0.95 | 0.93 | 0.99 | 0.91 | 0.81 | 0.91 | 0.98 | 1.02 | 0.89 | 0.81 | 0.95 | 1.01 | 0.98 | 0.95 |
| Rectum | 1.15 | 1.21 | 1.54 | 0.94 | 1.54 | 1.22 | 1.21 | 0.87 | 1.06 | 0.93 | 0.93 | 0.86 | 1.22 | 1.05 | 1.15 | 1.29 | 1.10 | 0.97 | 1.31 | 1.16 | 1.12 |
| Biliary tract | 0.74 | 0.43 | 0.71 | 0.74 | 0.68 | 0.59 | 0.68 | 0.61 | 0.55 | 0.55 | 0.59 | 0.64 | 0.73 | 0.67 | 0.61 | 0.73 | 0.75 | 0.71 | 0.76 | 0.75 | 0.67 |
| Gallbladder | 0.39 | 0.36 | 0.42 | 0.43 | 0.42 | 0.44 | 0.37 | 0.47 | 0.40 | 0.38 | 0.42 | 0.42 | 0.41 | 0.37 | 0.39 | 0.38 | 0.31 | 0.40 | 0.32 | 0.38 | 0.39 |

ASMR: Age-standardized mortality rate.

The distribution of ASMR by age group demonstrated that overall digestive system cancer mortality exceeded 1.00 starting from the 45–49 age group, after which it increased to 4.068 in the 70–74 age group, before gradually decreasing to 2.033 in the 85–89 age group. In the sex-stratified assessment, mortality exceeded 1.000 in men starting from the 50–54 age group. Gallbladder cancer showed earlier mortality peaks compared to liver and bile duct cancers (Table 5).

**Table 5.**
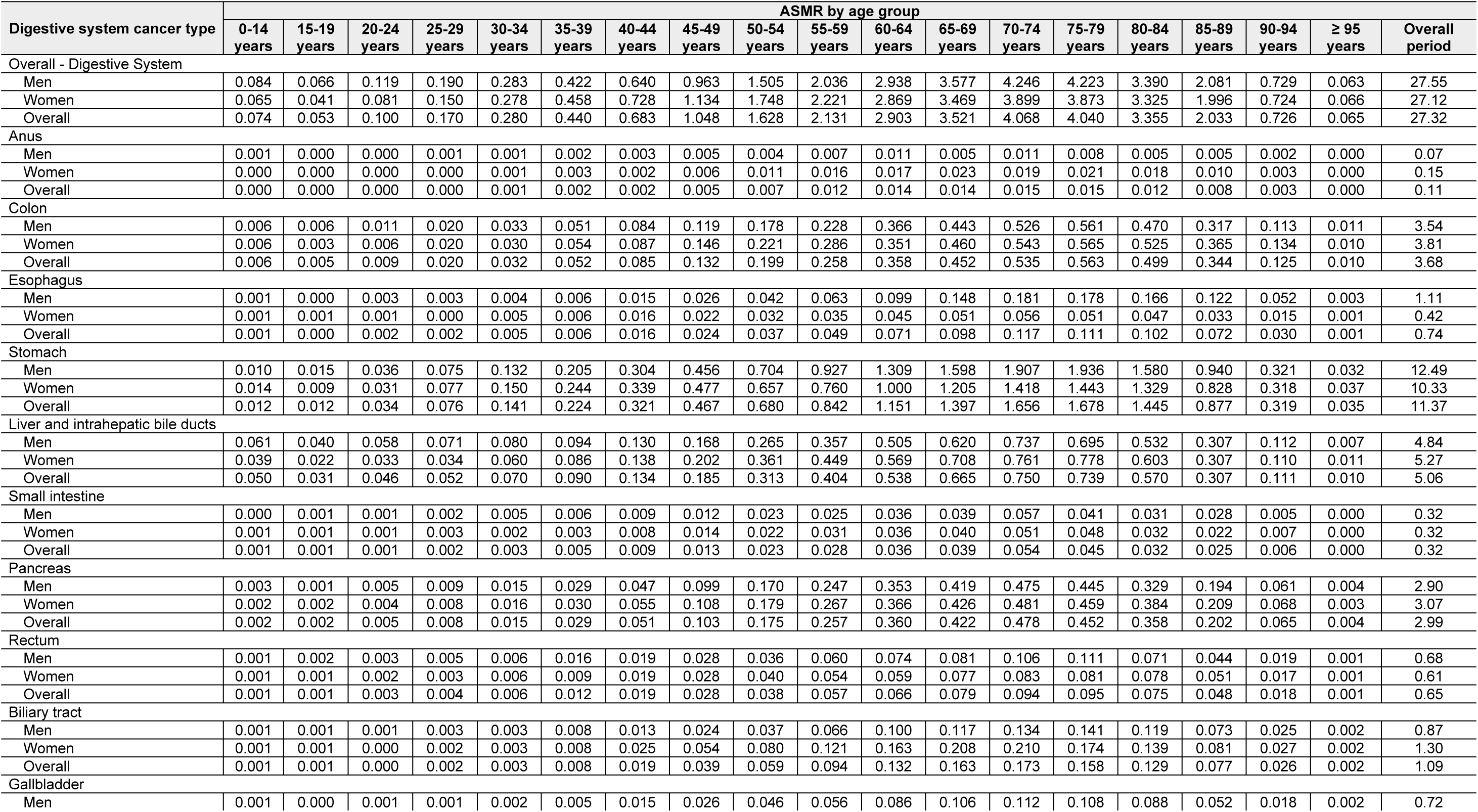
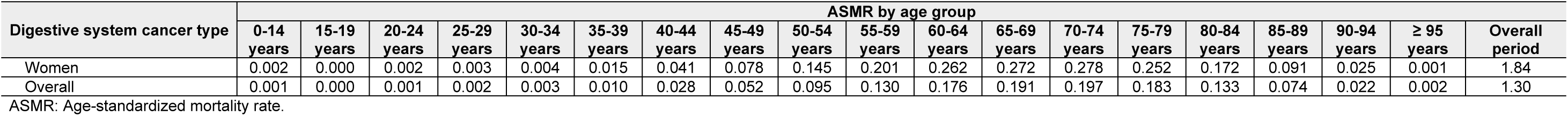
ASMR by age group, sex, and digestive system cancer type.

In the analysis by sex, the male-to-female ASMR ratio for esophageal cancer was found to be notably high, reaching 3.65 among those aged 85–89 and 4.04 among those aged 95 years and older, indicating a strong male predominance. Conversely, gallbladder cancer consistently showed higher mortality in women across almost all age groups, with the lowest sex ratios observed in the 25–29 (0.22) and 55–59 (0.28) age groups. (Table 6).

**Table 6.** ASMR sex ratio by age group and digestive system cancer type.

| Digestive system cancer type | ASMR sex ratio by age group |  |  |  |  |  |  |  |  |  |  |  |  |  |  |  |  |  | Overall period |
| --- | --- | --- | --- | --- | --- | --- | --- | --- | --- | --- | --- | --- | --- | --- | --- | --- | --- | --- | --- |
|  | 0-14 years | 15-19 years | 20-24 years | 25-29 years | 30-34 years | 35-39 years | 40-44 years | 45-49 years | 50-54 years | 55-59 years | 60-64 years | 65-69 years | 70-74 years | 75-79 years | 80-84 years | 85-89 years | 90-94 years | ≥ 95 years |  |
| Overall - Digestive System | 1.29 | 1.60 | 1.48 | 1.27 | 1.02 | 0.92 | 0.88 | 0.85 | 0.86 | 0.92 | 1.02 | 1.03 | 1.09 | 1.09 | 1.02 | 1.04 | 1.01 | 0.95 | 1.02 |
| Anus | 1.00 | 0.00 | 1.00 | 2.10 | 1.32 | 0.79 | 1.42 | 0.85 | 0.37 | 0.41 | 0.61 | 0.24 | 0.59 | 0.37 | 0.26 | 0.47 | 0.62 | 0.55 | 0.46 |
| Colon | 1.03 | 1.76 | 1.63 | 1.02 | 1.07 | 0.94 | 0.96 | 0.81 | 0.81 | 0.80 | 1.04 | 0.96 | 0.97 | 0.99 | 0.90 | 0.87 | 0.84 | 1.06 | 0.93 |
| Esophagus | 0.98 | 0.54 | 3.28 | 8.94 | 0.99 | 1.03 | 0.97 | 1.17 | 1.32 | 1.79 | 2.20 | 2.93 | 3.26 | 3.46 | 3.54 | 3.65 | 3.50 | 4.04 | 2.67 |
| Stomach | 0.74 | 1.57 | 1.15 | 0.98 | 0.88 | 0.84 | 0.90 | 0.96 | 1.07 | 1.22 | 1.31 | 1.33 | 1.34 | 1.34 | 1.19 | 1.13 | 1.01 | 0.86 | 1.21 |
| Liver and intrahepatic bile ducts | 1.58 | 1.81 | 1.77 | 2.10 | 1.33 | 1.08 | 0.95 | 0.83 | 0.73 | 0.80 | 0.89 | 0.88 | 0.97 | 0.89 | 0.88 | 1.00 | 1.01 | 0.69 | 0.92 |
| Small intestine | 0.31 | 0.50 | 1.33 | 0.75 | 2.40 | 1.96 | 1.12 | 0.90 | 1.04 | 0.83 | 0.99 | 0.98 | 1.12 | 0.85 | 0.98 | 1.27 | 0.70 | 0.91 | 1.00 |
| Pancreas | 1.45 | 0.37 | 1.15 | 1.08 | 0.99 | 0.97 | 0.86 | 0.92 | 0.95 | 0.93 | 0.97 | 0.98 | 0.99 | 0.97 | 0.86 | 0.93 | 0.90 | 1.32 | 0.95 |
| Rectum | 1.02 | 3.43 | 1.86 | 1.68 | 1.01 | 1.78 | 1.04 | 1.01 | 0.90 | 1.11 | 1.24 | 1.05 | 1.27 | 1.37 | 0.91 | 0.87 | 1.12 | 0.77 | 1.12 |
| Biliary tract | 2.01 | 1.04 | 1.96 | 1.79 | 0.99 | 1.07 | 0.52 | 0.44 | 0.46 | 0.54 | 0.61 | 0.56 | 0.64 | 0.81 | 0.85 | 0.90 | 0.95 | 1.43 | 0.67 |
| Gallbladder | 0.33 | 1.00 | 0.61 | 0.22 | 0.47 | 0.33 | 0.36 | 0.34 | 0.32 | 0.28 | 0.33 | 0.39 | 0.40 | 0.43 | 0.51 | 0.57 | 0.72 | 1.64 | 0.39 |

ASMR: Age-standardized mortality rate.

Furthermore, a female predominance in mortality was also evident for anal cancer and biliary tract cancer, which presented overall period ratios of 0.46 and 0.67, respectively. In contrast, stomach and rectal cancers exhibited an overall tendency towards higher mortality in men, with ratios of 1.21 and 1.12. Notably, for the digestive system overall, the sex ratio dropped below 1.00 between the ages of 35 and 59, reflecting higher mortality rates in women during these mid-life years. (Table 6).

Comparison of mortality between the period endpoints evidenced that, overall, there was a 0.1% reduction in ASMR for digestive system cancer between 2001 and 2020. However, in a stratification by sex, a 1.5% increase was found in men, while a reduction of the same magnitude occurred in women. In a differentiated analysis by cancer type, ASMR decreased for esophageal, stomach, and liver and bile duct cancers. Regarding the ranking by cancer type with the highest ASMR, stomach cancer ranked first, followed by colon cancer, with anal cancer in the last position. Comparatively between the period endpoints, colon cancer rose in the overall ASMR ranking, while liver and intrahepatic bile duct cancer dropped (Table 7).

**Table 7.**
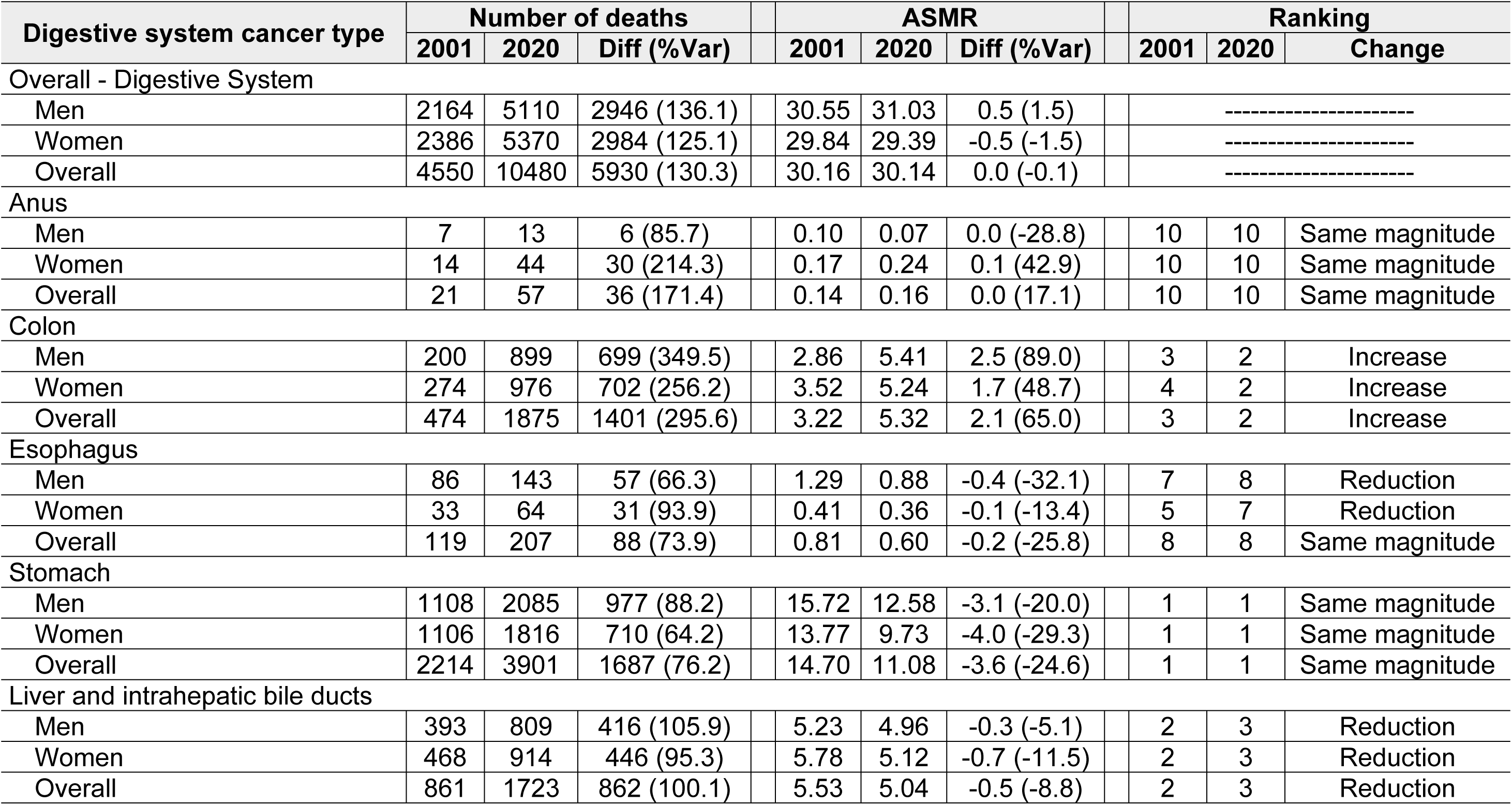
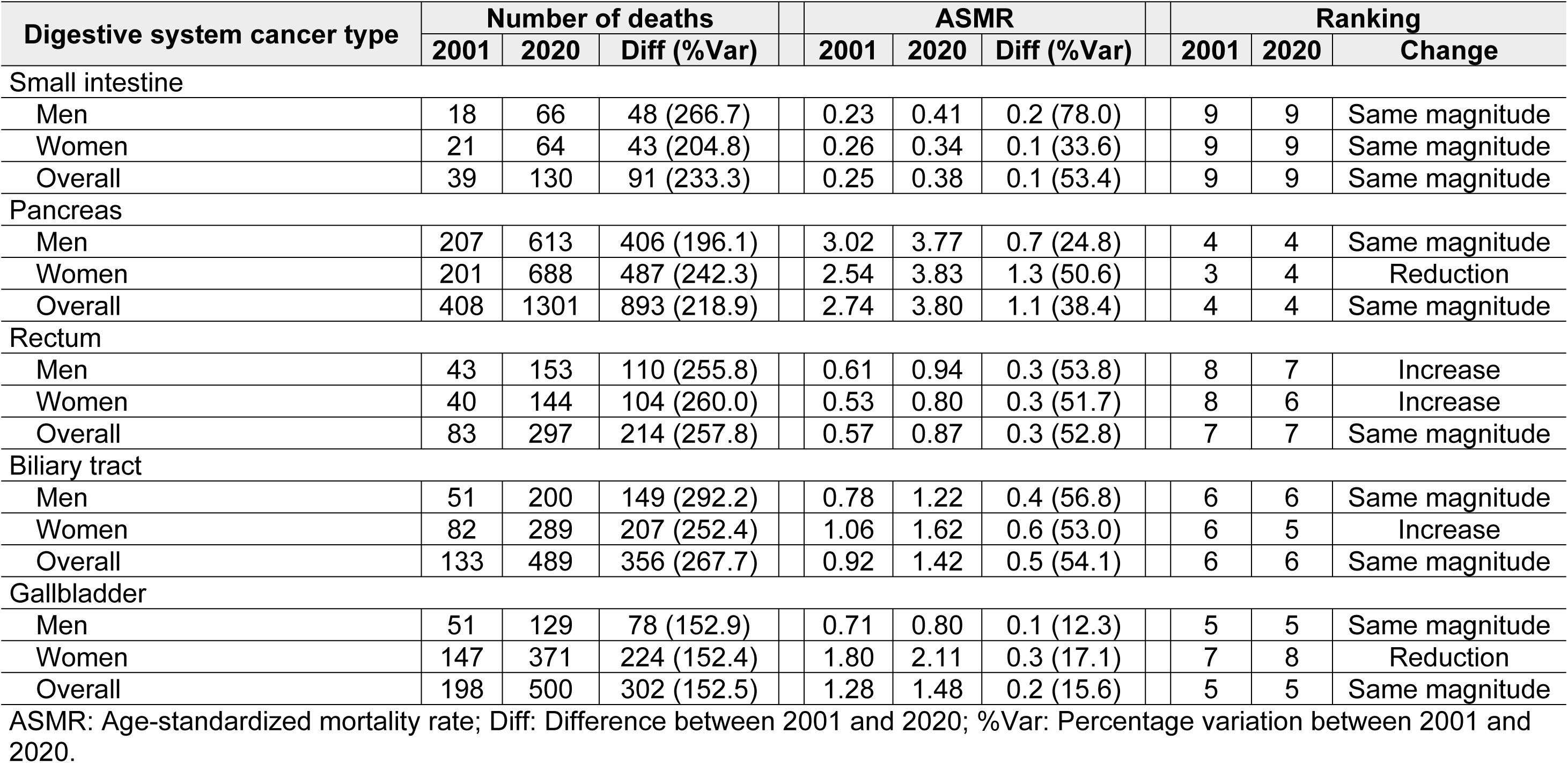
Mortality patterns by digestive system cancer type.

| Digestive system cancer type | Number of deaths |  |  | ASMR |  |  | Ranking |  |  |
| --- | --- | --- | --- | --- | --- | --- | --- | --- | --- |
|  | 2001 | 2020 | Diff (%Var) | 2001 | 2020 | Diff (%Var) | 2001 | 2020 | Change |
| Overall - Digestive System |  |  |  |  |  |  |  |  |  |
| Men | 2164 | 5110 | 2946 (136.1) | 30.55 | 31.03 | 0.5 (1.5) |  | ----- |  |
| Women | 2386 | 5370 | 2984 (125.1) | 29.84 | 29.39 | -0.5 (-1.5) |  | ----- |  |
| Overall | 4550 | 10480 | 5930 (130.3) | 30.16 | 30.14 | 0.0 (-0.1) |  | ----- |  |
| Anus |  |  |  |  |  |  |  |  |  |
| Men | 7 | 13 | 6 (85.7) | 0.10 | 0.07 | 0.0 (-28.8) | 10 | 10 | Same magnitude |
| Women | 14 | 44 | 30 (214.3) | 0.17 | 0.24 | 0.1 (42.9) | 10 | 10 | Same magnitude |
| Overall | 21 | 57 | 36 (171.4) | 0.14 | 0.16 | 0.0 (17.1) | 10 | 10 | Same magnitude |
| Colon |  |  |  |  |  |  |  |  |  |
| Men | 200 | 899 | 699 (349.5) | 2.86 | 5.41 | 2.5 (89.0) | 3 | 2 | Increase |
| Women | 274 | 976 | 702 (256.2) | 3.52 | 5.24 | 1.7 (48.7) | 4 | 2 | Increase |
| Overall | 474 | 1875 | 1401 (295.6) | 3.22 | 5.32 | 2.1 (65.0) | 3 | 2 | Increase |
| Esophagus |  |  |  |  |  |  |  |  |  |
| Men | 86 | 143 | 57 (66.3) | 1.29 | 0.88 | -0.4 (-32.1) | 7 | 8 | Reduction |
| Women | 33 | 64 | 31 (93.9) | 0.41 | 0.36 | -0.1 (-13.4) | 5 | 7 | Reduction |
| Overall | 119 | 207 | 88 (73.9) | 0.81 | 0.60 | -0.2 (-25.8) | 8 | 8 | Same magnitude |
| Stomach |  |  |  |  |  |  |  |  |  |
| Men | 1108 | 2085 | 977 (88.2) | 15.72 | 12.58 | -3.1 (-20.0) | 1 | 1 | Same magnitude |
| Women | 1106 | 1816 | 710 (64.2) | 13.77 | 9.73 | -4.0 (-29.3) | 1 | 1 | Same magnitude |
| Overall | 2214 | 3901 | 1687 (76.2) | 14.70 | 11.08 | -3.6 (-24.6) | 1 | 1 | Same magnitude |
| Liver and intrahepatic bile ducts |  |  |  |  |  |  |  |  |  |
| Men | 393 | 809 | 416 (105.9) | 5.23 | 4.96 | -0.3 (-5.1) | 2 | 3 | Reduction |
| Women | 468 | 914 | 446 (95.3) | 5.78 | 5.12 | -0.7 (-11.5) | 2 | 3 | Reduction |
| Overall | 861 | 1723 | 862 (100.1) | 5.53 | 5.04 | -0.5 (-8.8) | 2 | 3 | Reduction |

| Digestive system cancer type | Number of deaths |  |  |  | ASMR |  |  |  | Ranking |  |  |
| --- | --- | --- | --- | --- | --- | --- | --- | --- | --- | --- | --- |
|  | 2001 | 2020 | Diff (%Var) |  | 2001 | 2020 | Diff (%Var) |  | 2001 | 2020 | Change |
| Small intestine |  |  |  |  |  |  |  |  |  |  |  |
| Men | 18 | 66 | 48 (266.7) |  | 0.23 | 0.41 | 0.2 (78.0) |  | 9 | 9 | Same magnitude |
| Women | 21 | 64 | 43 (204.8) |  | 0.26 | 0.34 | 0.1 (33.6) |  | 9 | 9 | Same magnitude |
| Overall | 39 | 130 | 91 (233.3) |  | 0.25 | 0.38 | 0.1 (53.4) |  | 9 | 9 | Same magnitude |
| Pancreas |  |  |  |  |  |  |  |  |  |  |  |
| Men | 207 | 613 | 406 (196.1) |  | 3.02 | 3.77 | 0.7 (24.8) |  | 4 | 4 | Same magnitude |
| Women | 201 | 688 | 487 (242.3) |  | 2.54 | 3.83 | 1.3 (50.6) |  | 3 | 4 | Reduction |
| Overall | 408 | 1301 | 893 (218.9) |  | 2.74 | 3.80 | 1.1 (38.4) |  | 4 | 4 | Same magnitude |
| Rectum |  |  |  |  |  |  |  |  |  |  |  |
| Men | 43 | 153 | 110 (255.8) |  | 0.61 | 0.94 | 0.3 (53.8) |  | 8 | 7 | Increase |
| Women | 40 | 144 | 104 (260.0) |  | 0.53 | 0.80 | 0.3 (51.7) |  | 8 | 6 | Increase |
| Overall | 83 | 297 | 214 (257.8) |  | 0.57 | 0.87 | 0.3 (52.8) |  | 7 | 7 | Same magnitude |
| Biliary tract |  |  |  |  |  |  |  |  |  |  |  |
| Men | 51 | 200 | 149 (292.2) |  | 0.78 | 1.22 | 0.4 (56.8) |  | 6 | 6 | Same magnitude |
| Women | 82 | 289 | 207 (252.4) |  | 1.06 | 1.62 | 0.6 (53.0) |  | 6 | 5 | Increase |
| Overall | 133 | 489 | 356 (267.7) |  | 0.92 | 1.42 | 0.5 (54.1) |  | 6 | 6 | Same magnitude |
| Gallbladder |  |  |  |  |  |  |  |  |  |  |  |
| Men | 51 | 129 | 78 (152.9) |  | 0.71 | 0.80 | 0.1 (12.3) |  | 5 | 5 | Same magnitude |
| Women | 147 | 371 | 224 (152.4) |  | 1.80 | 2.11 | 0.3 (17.1) |  | 7 | 8 | Reduction |
| Overall | 198 | 500 | 302 (152.5) |  | 1.28 | 1.48 | 0.2 (15.6) |  | 5 | 5 | Same magnitude |

ASMR: Age-standardized mortality rate; Diff: Difference between 2001 and 2020; %Var: Percentage variation between 2001 and 2020.

The overall ASMR trend for the digestive system presented two segments: the first showed a significant reduction between 2001 and 2017 with an APC of -1.9 (95% CI: -2.5; -1.4), followed by a significant increase between 2017 and 2020 with an APC of 9.9 (95% CI: 3.6; 19.5). This pattern was similar between sexes for the first segment, while in the second segment, it was higher in men. Overall, colon, liver and intrahepatic bile ducts, pancreas, and gallbladder cancers presented three segments, where the last segment coincided in all cases between 2016 and 2020. Esophageal, small intestine, and biliary tract cancers presented a single segment throughout the period, where only esophageal cancer showed a downward trend. The number of segments varied by sex for liver and intrahepatic bile ducts, small intestine, pancreas, rectal, and biliary tract cancers (Table 8).

**Table 8.**
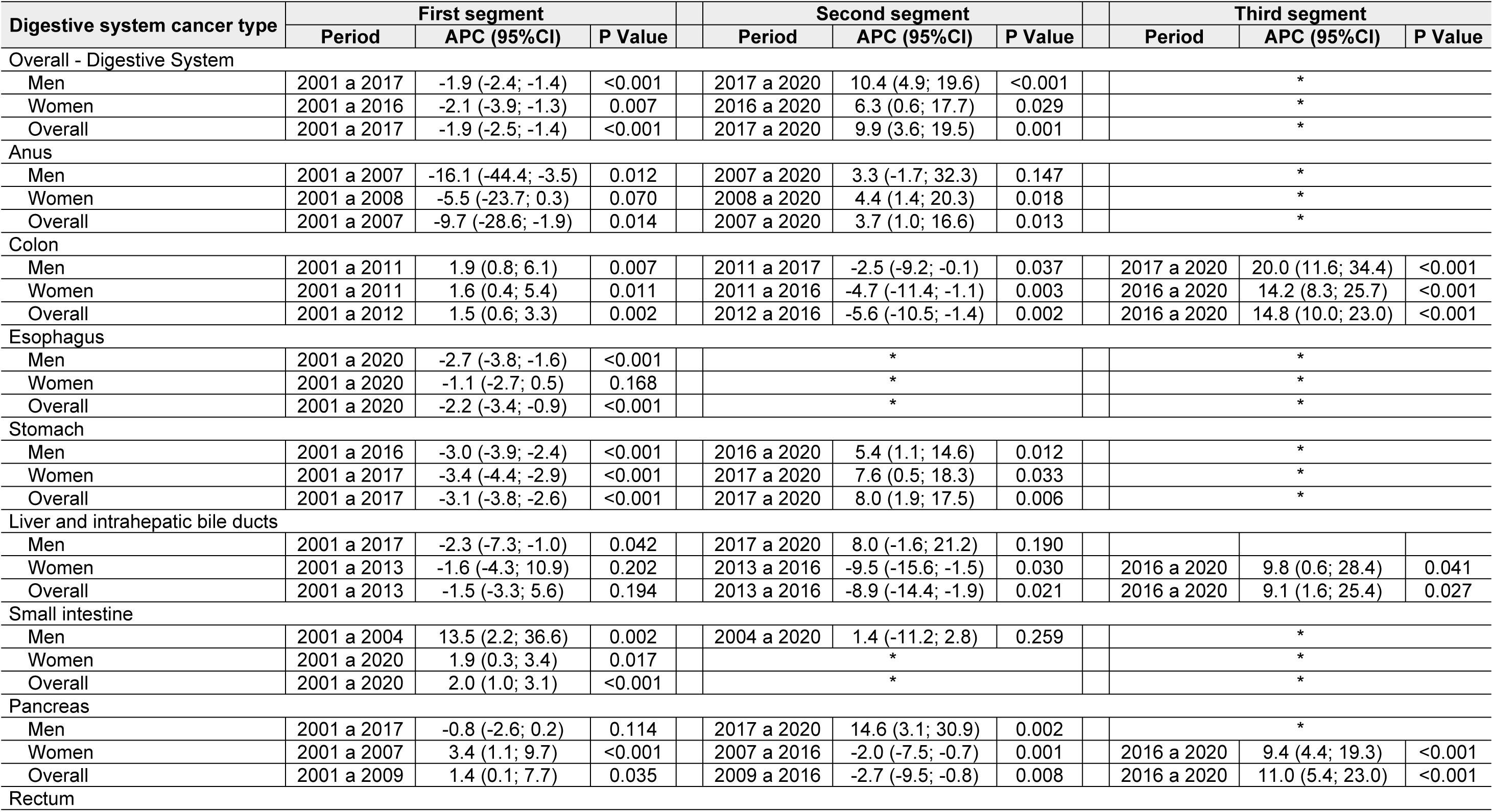
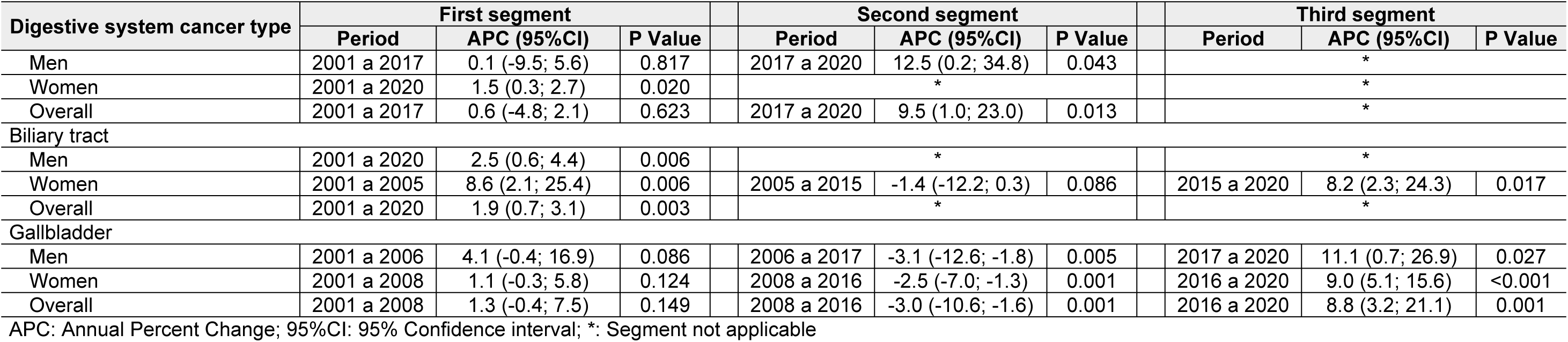
ASMR trends during the 2001–2020 period by sex and digestive system cancer type.

| Digestive system cancer type | First segment |  |  | Second segment |  |  | Third segment |  |  |
| --- | --- | --- | --- | --- | --- | --- | --- | --- | --- |
|  | Period | APC (95%CI) | P Value | Period | APC (95%CI) | P Value | Period | APC (95%CI) | P Value |
| Overall - Digestive System |  |  |  |  |  |  |  |  |  |
| Men | 2001 a 2017 | -1.9 (-2.4; -1.4) | <0.001 | 2017 a 2020 | 10.4 (4.9; 19.6) | <0.001 |  | * |  |
| Women | 2001 a 2016 | -2.1 (-3.9; -1.3) | 0.007 | 2016 a 2020 | 6.3 (0.6; 17.7) | 0.029 |  | * |  |
| Overall | 2001 a 2017 | -1.9 (-2.5; -1.4) | <0.001 | 2017 a 2020 | 9.9 (3.6; 19.5) | 0.001 |  | * |  |
| Anus |  |  |  |  |  |  |  |  |  |
| Men | 2001 a 2007 | -16.1 (-44.4; -3.5) | 0.012 | 2007 a 2020 | 3.3 (-1.7; 32.3) | 0.147 |  | * |  |
| Women | 2001 a 2008 | -5.5 (-23.7; 0.3) | 0.070 | 2008 a 2020 | 4.4 (1.4; 20.3) | 0.018 |  | * |  |
| Overall | 2001 a 2007 | -9.7 (-28.6; -1.9) | 0.014 | 2007 a 2020 | 3.7 (1.0; 16.6) | 0.013 |  | * |  |
| Colon |  |  |  |  |  |  |  |  |  |
| Men | 2001 a 2011 | 1.9 (0.8; 6.1) | 0.007 | 2011 a 2017 | -2.5 (-9.2; -0.1) | 0.037 | 2017 a 2020 | 20.0 (11.6; 34.4) | <0.001 |
| Women | 2001 a 2011 | 1.6 (0.4; 5.4) | 0.011 | 2011 a 2016 | -4.7 (-11.4; -1.1) | 0.003 | 2016 a 2020 | 14.2 (8.3; 25.7) | <0.001 |
| Overall | 2001 a 2012 | 1.5 (0.6; 3.3) | 0.002 | 2012 a 2016 | -5.6 (-10.5; -1.4) | 0.002 | 2016 a 2020 | 14.8 (10.0; 23.0) | <0.001 |
| Esophagus |  |  |  |  |  |  |  |  |  |
| Men | 2001 a 2020 | -2.7 (-3.8; -1.6) | <0.001 |  | * |  |  | * |  |
| Women | 2001 a 2020 | -1.1 (-2.7; 0.5) | 0.168 |  | * |  |  | * |  |
| Overall | 2001 a 2020 | -2.2 (-3.4; -0.9) | <0.001 |  | * |  |  | * |  |
| Stomach |  |  |  |  |  |  |  |  |  |
| Men | 2001 a 2016 | -3.0 (-3.9; -2.4) | <0.001 | 2016 a 2020 | 5.4 (1.1; 14.6) | 0.012 |  | * |  |
| Women | 2001 a 2017 | -3.4 (-4.4; -2.9) | <0.001 | 2017 a 2020 | 7.6 (0.5; 18.3) | 0.033 |  | * |  |
| Overall | 2001 a 2017 | -3.1 (-3.8; -2.6) | <0.001 | 2017 a 2020 | 8.0 (1.9; 17.5) | 0.006 |  | * |  |
| Liver and intrahepatic bile ducts |  |  |  |  |  |  |  |  |  |
| Men | 2001 a 2017 | -2.3 (-7.3; -1.0) | 0.042 | 2017 a 2020 | 8.0 (-1.6; 21.2) | 0.190 |  |  |  |
| Women | 2001 a 2013 | -1.6 (-4.3; 10.9) | 0.202 | 2013 a 2016 | -9.5 (-15.6; -1.5) | 0.030 | 2016 a 2020 | 9.8 (0.6; 28.4) | 0.041 |
| Overall | 2001 a 2013 | -1.5 (-3.3; 5.6) | 0.194 | 2013 a 2016 | -8.9 (-14.4; -1.9) | 0.021 | 2016 a 2020 | 9.1 (1.6; 25.4) | 0.027 |
| Small intestine |  |  |  |  |  |  |  |  |  |
| Men | 2001 a 2004 | 13.5 (2.2; 36.6) | 0.002 | 2004 a 2020 | 1.4 (-11.2; 2.8) | 0.259 |  | * |  |
| Women | 2001 a 2020 | 1.9 (0.3; 3.4) | 0.017 |  | * |  |  | * |  |
| Overall | 2001 a 2020 | 2.0 (1.0; 3.1) | <0.001 |  | * |  |  | * |  |
| Pancreas |  |  |  |  |  |  |  |  |  |
| Men | 2001 a 2017 | -0.8 (-2.6; 0.2) | 0.114 | 2017 a 2020 | 14.6 (3.1; 30.9) | 0.002 |  | * |  |
| Women | 2001 a 2007 | 3.4 (1.1; 9.7) | <0.001 | 2007 a 2016 | -2.0 (-7.5; -0.7) | 0.001 | 2016 a 2020 | 9.4 (4.4; 19.3) | <0.001 |
| Overall | 2001 a 2009 | 1.4 (0.1; 7.7) | 0.035 | 2009 a 2016 | -2.7 (-9.5; -0.8) | 0.008 | 2016 a 2020 | 11.0 (5.4; 23.0) | <0.001 |
| Rectum |  |  |  |  |  |  |  |  |  |

| Digestive system cancer type | First segment |  |  |  | Second segment |  |  |  | Third segment |  |  |
| --- | --- | --- | --- | --- | --- | --- | --- | --- | --- | --- | --- |
|  | Period | APC (95%CI) | P Value |  | Period | APC (95%CI) | P Value |  | Period | APC (95%CI) | P Value |
| Men | 2001 a 2017 | 0.1 (-9.5; 5.6) | 0.817 |  | 2017 a 2020 | 12.5 (0.2; 34.8) | 0.043 |  |  | * |  |
| Women | 2001 a 2020 | 1.5 (0.3; 2.7) | 0.020 |  |  | * |  |  |  | * |  |
| Overall | 2001 a 2017 | 0.6 (-4.8; 2.1) | 0.623 |  | 2017 a 2020 | 9.5 (1.0; 23.0) | 0.013 |  |  | * |  |
| Biliary tract |  |  |  |  |  |  |  |  |  |  |  |
| Men | 2001 a 2020 | 2.5 (0.6; 4.4) | 0.006 |  |  | * |  |  |  | * |  |
| Women | 2001 a 2005 | 8.6 (2.1; 25.4) | 0.006 |  | 2005 a 2015 | -1.4 (-12.2; 0.3) | 0.086 |  | 2015 a 2020 | 8.2 (2.3; 24.3) | 0.017 |
| Overall | 2001 a 2020 | 1.9 (0.7; 3.1) | 0.003 |  |  | * |  |  |  | * |  |
| Gallbladder |  |  |  |  |  |  |  |  |  |  |  |
| Men | 2001 a 2006 | 4.1 (-0.4; 16.9) | 0.086 |  | 2006 a 2017 | -3.1 (-12.6; -1.8) | 0.005 |  | 2017 a 2020 | 11.1 (0.7; 26.9) | 0.027 |
| Women | 2001 a 2008 | 1.1 (-0.3; 5.8) | 0.124 |  | 2008 a 2016 | -2.5 (-7.0; -1.3) | 0.001 |  | 2016 a 2020 | 9.0 (5.1; 15.6) | <0.001 |
| Overall | 2001 a 2008 | 1.3 (-0.4; 7.5) | 0.149 |  | 2008 a 2016 | -3.0 (-10.6; -1.6) | 0.001 |  | 2016 a 2020 | 8.8 (3.2; 21.1) | 0.001 |

APC: Annual Percent Change; 95%CI: 95% Confidence interval; \*: Segment not applicable

For ASMR projection to 2030, two predicted periods were estimated: the first from 2021 to 2025, and the second from 2026 to 2030. Generally, for digestive system cancers, it was found that in the first predicted period (2021–2025), the ASMR would be lower compared to the previous period (2016–2020), with a lower value in women, followed by an increase in the second predicted period (2026–2030). The ASMR for the last predicted period was lower than the last observed period (2016–2020) for esophageal and liver and bile duct cancers in both men and women, whereas for stomach cancer, this reduction would occur only in women. Regarding cancers maintaining the same value in the last predicted period compared to the last observed period, this was identified for anal, small intestine, and gallbladder cancers (Table 9).

**Table 9.**
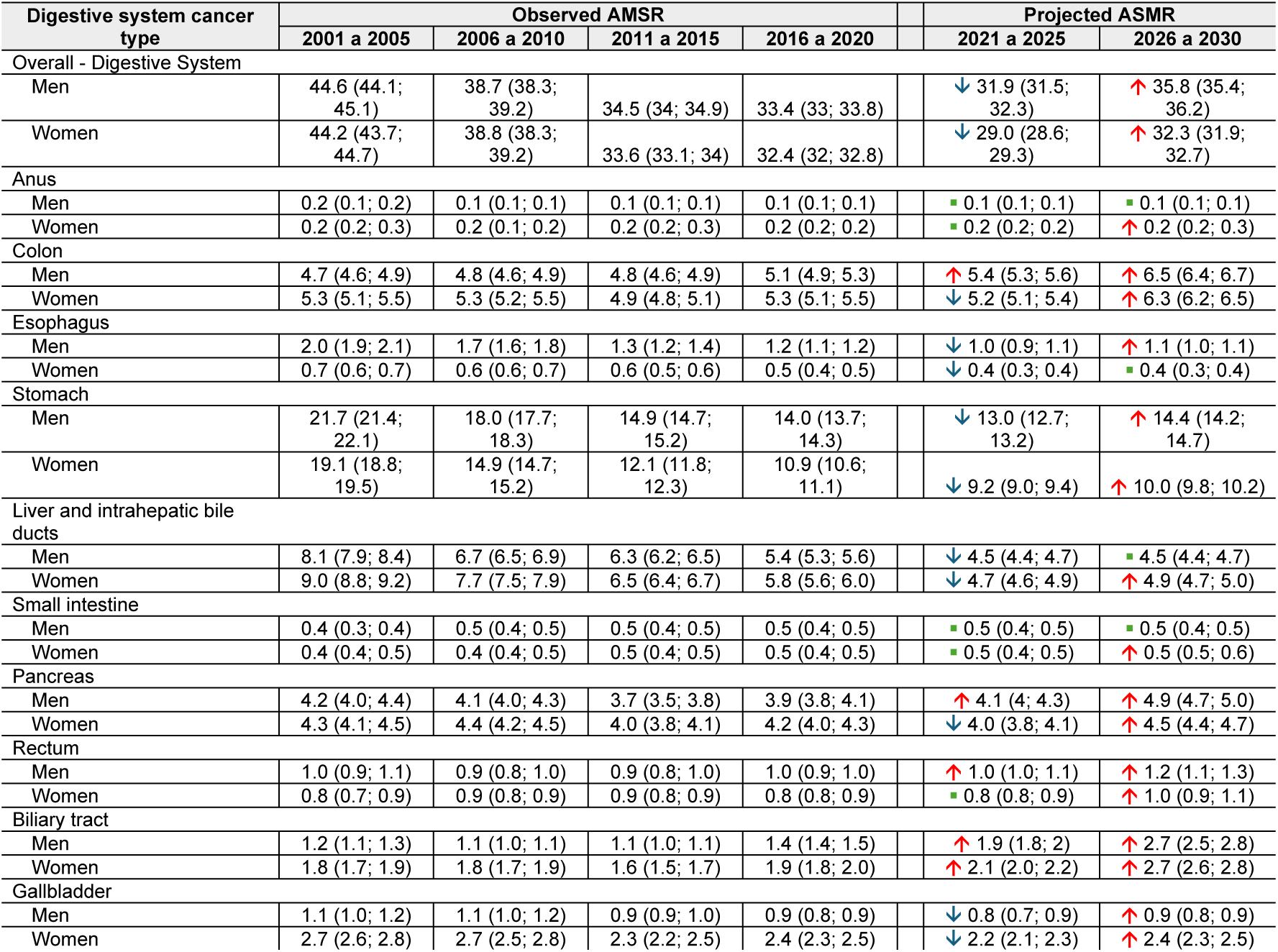
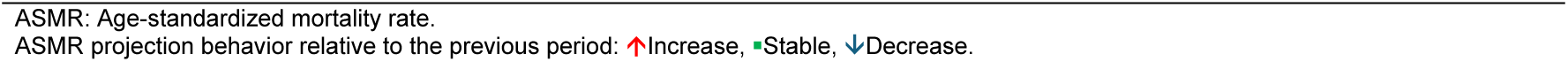
ASMR projection to 2030 by digestive system cancer type and sex.

| Digestive system cancer type | Observed ASMR |  |  |  | Projected ASMR |  |
| --- | --- | --- | --- | --- | --- | --- |
|  | 2001 a 2005 | 2006 a 2010 | 2011 a 2015 | 2016 a 2020 | 2021 a 2025 | 2026 a 2030 |
| Overall - Digestive System |  |  |  |  |  |  |
| Men | 44.6 (44.1; 45.1) | 38.7 (38.3; 39.2) | 34.5 (34; 34.9) | 33.4 (33; 33.8) | ↓ 31.9 (31.5; 32.3) | ↑ 35.8 (35.4; 36.2) |
| Women | 44.2 (43.7; 44.7) | 38.8 (38.3; 39.2) | 33.6 (33.1; 34) | 32.4 (32; 32.8) | ↓ 29.0 (28.6; 29.3) | ↑ 32.3 (31.9; 32.7) |
| Anus |  |  |  |  |  |  |
| Men | 0.2 (0.1; 0.2) | 0.1 (0.1; 0.1) | 0.1 (0.1; 0.1) | 0.1 (0.1; 0.1) | ■ 0.1 (0.1; 0.1) | ■ 0.1 (0.1; 0.1) |
| Women | 0.2 (0.2; 0.3) | 0.2 (0.1; 0.2) | 0.2 (0.2; 0.3) | 0.2 (0.2; 0.2) | ■ 0.2 (0.2; 0.2) | ↑ 0.2 (0.2; 0.3) |
| Colon |  |  |  |  |  |  |
| Men | 4.7 (4.6; 4.9) | 4.8 (4.6; 4.9) | 4.8 (4.6; 4.9) | 5.1 (4.9; 5.3) | ↑ 5.4 (5.3; 5.6) | ↑ 6.5 (6.4; 6.7) |
| Women | 5.3 (5.1; 5.5) | 5.3 (5.2; 5.5) | 4.9 (4.8; 5.1) | 5.3 (5.1; 5.5) | ↓ 5.2 (5.1; 5.4) | ↑ 6.3 (6.2; 6.5) |
| Esophagus |  |  |  |  |  |  |
| Men | 2.0 (1.9; 2.1) | 1.7 (1.6; 1.8) | 1.3 (1.2; 1.4) | 1.2 (1.1; 1.2) | ↓ 1.0 (0.9; 1.1) | ↑ 1.1 (1.0; 1.1) |
| Women | 0.7 (0.6; 0.7) | 0.6 (0.6; 0.7) | 0.6 (0.5; 0.6) | 0.5 (0.4; 0.5) | ↓ 0.4 (0.3; 0.4) | ■ 0.4 (0.3; 0.4) |
| Stomach |  |  |  |  |  |  |
| Men | 21.7 (21.4; 22.1) | 18.0 (17.7; 18.3) | 14.9 (14.7; 15.2) | 14.0 (13.7; 14.3) | ↓ 13.0 (12.7; 13.2) | ↑ 14.4 (14.2; 14.7) |
| Women | 19.1 (18.8; 19.5) | 14.9 (14.7; 15.2) | 12.1 (11.8; 12.3) | 10.9 (10.6; 11.1) | ↓ 9.2 (9.0; 9.4) | ↑ 10.0 (9.8; 10.2) |
| Liver and intrahepatic bile ducts |  |  |  |  |  |  |
| Men | 8.1 (7.9; 8.4) | 6.7 (6.5; 6.9) | 6.3 (6.2; 6.5) | 5.4 (5.3; 5.6) | ↓ 4.5 (4.4; 4.7) | ■ 4.5 (4.4; 4.7) |
| Women | 9.0 (8.8; 9.2) | 7.7 (7.5; 7.9) | 6.5 (6.4; 6.7) | 5.8 (5.6; 6.0) | ↓ 4.7 (4.6; 4.9) | ↑ 4.9 (4.7; 5.0) |
| Small intestine |  |  |  |  |  |  |
| Men | 0.4 (0.3; 0.4) | 0.5 (0.4; 0.5) | 0.5 (0.4; 0.5) | 0.5 (0.4; 0.5) | ■ 0.5 (0.4; 0.5) | ■ 0.5 (0.4; 0.5) |
| Women | 0.4 (0.4; 0.5) | 0.4 (0.4; 0.5) | 0.5 (0.4; 0.5) | 0.5 (0.4; 0.5) | ■ 0.5 (0.4; 0.5) | ↑ 0.5 (0.5; 0.6) |
| Pancreas |  |  |  |  |  |  |
| Men | 4.2 (4.0; 4.4) | 4.1 (4.0; 4.3) | 3.7 (3.5; 3.8) | 3.9 (3.8; 4.1) | ↑ 4.1 (4; 4.3) | ↑ 4.9 (4.7; 5.0) |
| Women | 4.3 (4.1; 4.5) | 4.4 (4.2; 4.5) | 4.0 (3.8; 4.1) | 4.2 (4.0; 4.3) | ↓ 4.0 (3.8; 4.1) | ↑ 4.5 (4.4; 4.7) |
| Rectum |  |  |  |  |  |  |
| Men | 1.0 (0.9; 1.1) | 0.9 (0.8; 1.0) | 0.9 (0.8; 1.0) | 1.0 (0.9; 1.0) | ↑ 1.0 (1.0; 1.1) | ↑ 1.2 (1.1; 1.3) |
| Women | 0.8 (0.7; 0.9) | 0.9 (0.8; 0.9) | 0.9 (0.8; 0.9) | 0.8 (0.8; 0.9) | ■ 0.8 (0.8; 0.9) | ↑ 1.0 (0.9; 1.1) |
| Biliary tract |  |  |  |  |  |  |
| Men | 1.2 (1.1; 1.3) | 1.1 (1.0; 1.1) | 1.1 (1.0; 1.1) | 1.4 (1.4; 1.5) | ↑ 1.9 (1.8; 2) | ↑ 2.7 (2.5; 2.8) |
| Women | 1.8 (1.7; 1.9) | 1.8 (1.7; 1.9) | 1.6 (1.5; 1.7) | 1.9 (1.8; 2.0) | ↑ 2.1 (2.0; 2.2) | ↑ 2.7 (2.6; 2.8) |
| Gallbladder |  |  |  |  |  |  |
| Men | 1.1 (1.0; 1.2) | 1.1 (1.0; 1.2) | 0.9 (0.9; 1.0) | 0.9 (0.8; 0.9) | ↓ 0.8 (0.7; 0.9) | ↑ 0.9 (0.8; 0.9) |
| Women | 2.7 (2.6; 2.8) | 2.7 (2.5; 2.8) | 2.3 (2.2; 2.5) | 2.4 (2.3; 2.5) | ↓ 2.2 (2.1; 2.3) | ↑ 2.4 (2.3; 2.5) |

## DISCUSSION

### Main findings

Digestive system cancer is a global public health concern affecting individuals of all ages, impairing quality of life due to its morbidity, and tending towards mortality as part of its pathophysiological evolution. In this context, the present study identified that mortality, evaluated via ASMR, generally showed a downward trend until 2016, followed by an increase. However, differentiated analysis by cancer type revealed that only esophageal cancer maintained a constant downward trend, while biliary tract cancer showed an increase. Projections for 2026–2030 evidenced that three (anus, small intestine, and gallbladder) of the ten digestive cancer types would maintain ASMR values similar to those of 2016–2020, while for two types (esophagus, and liver and intrahepatic bile ducts), the ASMR would be lower.

### Results in context

Our findings regarding ASMR patterns demonstrated important differences according to cancer type, which justifies differentiated evaluations. For instance, the Global Burden of Disease 2019 Cancer Collaboration presented a global report on ASMR by cancer type based on data from “The Global Burden of Diseases, Injuries, and Risk Factors Study 2019,” identifying colorectal cancer (ASMR=13.7) and stomach cancer (ASMR=11.9) among the top five causes of mortality in 2019 (15). These results resemble our data, where the ASMR for stomach cancer was 11.4, while for colorectal cancer (including anus) it was 4.4. The discrepancies between global data and our findings in Peru could be attributed to the specific conditions of our population regarding the distribution of risk factors and the characteristics of healthcare supply to mitigate mortality, mainly related to diagnosis at advanced stages and poor access to innovative treatments that have notably improved survival in other contexts.

A study published in 2016 evaluating changes in global ASMR between 2000 and 2010 identified a reduction in stomach cancer mortality for both men and women, finding that ASMR in men varied from 12.02 to 9.82, and in women from 5.76 to 4.76 (16). These results agree with our findings, which also showed a reduction in ASMR, although the magnitude of mortality in women was higher in our data. Differences in magnitude may be attributed to the fact that our data analyse a single country rather than global figures, where implemented health policies may impact mortality predominantly in women, who exhibit greater utilization of health services (17).

An assessment of stomach cancer mortality performed separately by sex in five countries (Japan, UK, USA, Italy, and France) using WHO mortality database records between 1950 and 2008 identified a downward trend in mortality, which was greater in men than in women (18). These results align with our study, where a reduction of approximately 3% was found between the start of the period (2001) and 2008, with ASMR also being higher in men. However, our study covers a more recent period where an increase in mortality was observed.

Concerning esophageal cancer, Qiu et al. (19), using WHO mortality database records between 1960 and 2000, identified a downward trend in mortality, reporting higher ASMR in men than in women. Comparing this with our results, we highlight the coincidence regarding the downward trend in ASMR, where we also found this measure to be higher in men; however, it is necessary to note that our data are more recent, beginning where the period reported by Qiu et al. ends.

In 2001, Barrios et al. evaluated esophageal cancer mortality in men and women, identifying an ASMR of 1.1 in men and 0.4 in women, with a ratio of 2.9 (20). These results agree with the ratio identified in our study, where the average ASMR for the period was 2.7.

When analyzing the gastroesophageal component, evidence shows that while both esophageal and stomach cancers exhibit a downward trend, this is sustained for the esophagus, whereas stomach cancer presents an increase in mortality in recent years. This fact could be due to aspects related to medication use (21), as well as Helicobacter pylori infection (22,23). Furthermore, it could be related to the continued diagnosis of patients with advanced cancer and the fact that, although the prognosis of these patients has improved globally thanks to the development of targeted therapy and immunotherapy, these treatments are not widely available in the national reality.

Mortality from colon, rectal, and anal cancer was higher in men across the different studies evaluated, notably tending to be lower in Asia, Europe, Australia, and the United States between 1990 and 2005 (24). In a broader study including Latin American countries, it was identified that by 2012, the ASMR for the last 10 years in Colombia was 1.9 for men and 0.9 for women (25).

Sierra et al. conducted a study identifying that the colorectal cancer ASMR in Peru between 2001 and 2005 was slightly higher in women (3.4 vs. 3.3 in men), with a sex ratio of 1.0 (26). In our research, we considered a longer study period as well as a broader age range, which could explain the differences found.

In Ireland, O’Lorcain et al. (27), using WHO mortality database records between 1950 and 2017, identified that colon cancer ASMR was higher in men than in women and that a downward trend existed from 1986 onwards. This partially resembles our results, where we found an irregular downward trend between 2001 and 2016 with higher values in women, followed by a marked increasing trend in ASMR for both sexes.

Luo et al. (7) evaluated mortality trends in Australia using data from the New South Wales Cancer Registry from 1972 to 2015, with a projection to 2040. They reported that at the beginning of the period, the overall ASMR was 11.6 (12.4 in men, 11.0 in women), dropping to 6.1, 7.0, and 5.3 respectively by 2015, evidencing a clear downward trend over time. Their estimation maintained this trend towards 2030 with values of 4.3, 5.0, and 3.7, respectively. The aforementioned results partially disagree with our projection findings, where we identified that in the first projection segment (2021–2025), a reduction would occur in both sexes (slightly more marked in women), followed by an increase in the 2026–2030 period, predominantly in men. In addition to the difference in country context, the discrepancy could be explained by differences in the data source, the observed data period used, and the predicted interval.

In the specific case of rectal cancer, we highlight the report by Luo et al. (7), who also evaluated mortality for this cancer type separately, finding it was higher in men and showed a downward pattern, which was more homogeneous in women. Their projection found that at the start of the observed period (1972), the overall ASMR was 5.5 (7.0 for men, 4.4 for women), figures that became 3.7, 4.9, and 2.6 respectively by 2015. Regarding the 2030 projection, these figures were 3.1, 4.0, and 2.3 in the same order. Regarding our results, we emphasize that changes in ASMR were not as homogeneous over time; likewise, we found an increasing trend in the predicted estimates for the 2021–2025 and 2026–2030 periods.

Regarding anal cancer, it should be noted that literature usually evaluates this structure together with the rectum and colon. However, our study conducted a differentiated analysis finding a distinct pattern by sex, where ASMR was higher in women than in men, presenting an irregular trend over time and a different projection by sex, with a slight reduction in women and an increase in men. These results could prompt reflection on the potential existence of other factors presenting differentially between men and women, such as Human Papillomavirus (HPV) infection (28), the presence of hemorrhoids (29), and other conditions.

For pancreatic cancer, Yin et al. (6) evaluated mortality records in China between 2006 and 2015 using the National Central Cancer Registry of China, finding an irregular increasing pattern in ASMR, ranging generally from 3.35 in 2006 to 3.78 in 2015, with a projection of 4.65 for 2025. By sex, these figures were 3.99, 4.50, and 5.58 in men, and 2.74, 3.07, and 3.62 in women, respectively. Compared with our results, we highlight that figures of similar magnitude were presented in Peru, although higher mortality was found in women, with predicted ASMR values for the 2021–2025 period of 4.1 for men and 4.0 for women. Regarding the discrepancies found, these could be attributed to the number of years used as well as the statistical methods employed, as the projection by Yin et al. was performed using linear regression.

In a study by Jiang et al. (30) focusing on pancreatic cancer mortality in the Western Pacific region, ASMR was found to increase between 1990 and 2020, with higher values in men than in women. They also found that cases exceeded 5,000 in men starting at ages 40–44, while in women this occurred between 55–59, with a peak for both sexes between 70–74 years. Regarding these results, it is worth noting that in our study, ASMR was higher in women with a heterogeneous distribution over time; however, there is concordance regarding the age group presenting the highest mortality for this cancer type.

Regarding liver cancer mortality, Liu et al. (31) analyzed death records for this condition between 2008 and 2020 using the Disease Surveillance Points system of the Chinese Center for Disease Control and Prevention. They reported that ASMR was higher in men but that overall and in both sexes, there was a clear downward trend, reaching 2020 values of 19.92 overall, 30.15 in men, and 10.01 in women. The projection to 2030 identified a downward trend to 16.86 overall, 18.83 in men, and 5.60 in women. According to our results, liver cancer mortality in Peru over time was higher in women, and although a downward trend existed, it increased from 2016 to 2020; nevertheless, the 2030 projection suggests that ASMR will tend to reduce. It should be specified that the ASMR reported by Liu et al. was higher than that presented in Peru.

For gallbladder and biliary tract cancer, Chen et al. (5) analyzed ASMR trends in young adults aged 15–49 between 1990 and 2019, with a projection for 2020–2035 using Global Health Data Exchange records. They found that at the start of the period, the ASMR was 0.22 overall, 0.24 in men, and 0.20 in women, while for 2019 these figures were 0.30, 0.37, and 0.22 respectively; projection showed that ASMR would increase overall and in both sexes. In contrast, our results evidence that ASMR in Peru was higher in women and that the estimated projection contemplates a mortality reduction for the 2021–2025 and 2026–2030 periods; aspects that must be considered taking into account population differences, age restriction, and data origin.

According to the analyzed data, digestive system cancer ASMR peaks between 70 and 79 years, presenting early onset in stomach and rectal cancer, while late mortality is more present in liver and biliary tract cancer. This differentiation in age susceptibility for mortality may respond to the types of noxious agents affecting the organs and their respective mechanisms of action.

Observed ASMR trend changes in the study period allow us to identify that most cancer types exhibit more than one segment, where the second segment occurs between 2005–2008, 2011–2013, or 2015–2017. Likewise, in cancer types with three segments in their trend, the latter usually begins between 2015–2017. Regarding these figures, it is important to highlight that periods starting between 2015–2017 showed an increase in APC compared to previous segments.

To understand aspects potentially linked to our findings, it is important to comprehend that during the 2006–2011 period, the government focused efforts on prioritizing health promotion, albeit with scarce favorable results due to limited human and economic resources. Subsequently, from 2011 to 2016, a results-based budgeting approach was followed, where Universal Health Coverage (Aseguramiento Universal en Salud) was promoted under the financing of the Comprehensive Health Insurance (Sistema Integral de Salud - SIS) with active decentralized management (32).

It is in this scenario that “Plan Esperanza” was implemented in Peru, aiming to boost financing for cancer prevention and control (33), which had significant activity between 2012 and 2016, stages where our data found a reduction in ASMR. However, beyond economic, political, coverage, and healthcare access aspects, changes at the environmental and lifestyle levels must be considered.

### Strengths and limitations

The observed ASMR pattern is habitually analyzed using different existing databases; however, while knowing how health phenomena present is important, it is also relevant to identify how such phenomena might continue to present in future hypothetical scenarios. Thus, projection analyses gain significant value at the epidemiological level for health service supply planning. Through our research, we performed a predictive evaluation to estimate ASMR with a five-year horizon until 2030, a period also proposed by other studies.

Differences found in ASMR projection estimates may be due to data availability, as projection quality is affected by the number of observed moments or periods, how recent these are relative to the projection period, and the length of the predicted segment. Our research utilizes more recent data (year 2020) and requires a shorter projection segment (10 years).

While some studies in the scientific literature have performed trend and projection evaluations of cancer ASMR, our research differs by conducting a centralized approach on digestive system cancers, presenting a stratified analysis for each; additionally, we address the study problem from a descriptive, analytical, and predictive perspective, seeking to provide a robust contribution to the available evidence.

However, this research presents limitations inherent to the nature of the data used. As secondary data from an existing registry were used, a potential information bias due to underreporting could exist, since mortality information is reported by health professionals nationwide who attend these cases, implying possible human error at the time of registration. Nevertheless, it is important to specify that mortality records are the responsibility of the attending physician, who has knowledge of the patients’ pathologies.

Another potential limitation in the database concerns the possible loss of data regarding non-institutionalized deaths, where correct information registration was not possible. Notwithstanding, our findings seek to provide an overview of the dynamics of cancer mortality over time, as well as a hypothetical estimate of how it might present in the future.

### Conclusions

Digestive system cancer mortality is a phenomenon characterized by heterogeneous patterns, presenting variations according to time and cancer type. Most of these pathologies exhibited more than one trend throughout the study years; although there was generally a reduction in ASMR in most cases, an analysis of the final period segments evidences an increasing trend in ASMR. Regarding cancer mortality projections, it was evidenced that, compared to the last evaluated five-year period (2016–2020), the ASMR for the 2026–2030 period will be higher for most evaluated cancer types, although presenting with different magnitudes by sex.

## Author contributions

**Conceptualization:** Gilmer Solis-Sánchez, Oscar Augusto Lengua-Olivares, Maricela Curisinche-Rojas, Margot Haydee Vidal-Anzardo, Johanny Fidela de Fátima Muro-Cieza, Karina Mayra Aliaga-Llerena de Nuñez.

**Data curation:** Gilmer Solis-Sánchez, Oscar Augusto Lengua-Olivares.

**Formal analysis:** Gilmer Solis-Sánchez, Oscar Augusto Lengua-Olivares.

**Investigation:** Gilmer Solis-Sánchez, Oscar Augusto Lengua-Olivares, Margot Haydee Vidal-Anzardo, Johanny Fidela de Fátima Muro-Cieza, Karina Mayra Aliaga-Llerena de Nuñez.

**Methodology:** Gilmer Solis-Sánchez, Margot Haydee Vidal-Anzardo, Johanny Fidela de Fátima Muro-Cieza, Karina Mayra Aliaga-Llerena de Nuñez.

**Project administration:** Gilmer Solis-Sánchez, Maricela Curisinche-Rojas, Johanny Fidela de Fátima Muro-Cieza, Karina Mayra Aliaga-Llerena de Nuñez.

**Resources:** Maricela Curisinche-Rojas, Johanny Fidela de Fátima Muro-Cieza, Karina Mayra Aliaga-Llerena de Nuñez

**Software:** Gilmer Solis-Sánchez, Oscar Augusto Lengua-Olivares.

**Supervision:** Maricela Curisinche-Rojas, Margot Haydee Vidal-Anzardo, Johanny Fidela de Fátima Muro-Cieza, Karina Mayra Aliaga-Llerena de Nuñez.

**Validation:** Maricela Curisinche-Rojas, Margot Haydee Vidal-Anzardo, Johanny Fidela de Fátima Muro-Cieza, Karina Mayra Aliaga-Llerena de Nuñez.

**Visualization:** Gilmer Solis-Sánchez.

**Writing – original draft:** Gilmer Solis-Sánchez, Oscar Augusto Lengua-Olivares, Maricela Curisinche-Rojas, Margot Haydee Vidal-Anzardo, Johanny Fidela de Fátima Muro-Cieza, Karina Mayra Aliaga-Llerena de Nuñez.

**Writing – review & editing:** Gilmer Solis-Sánchez, Oscar Augusto Lengua-Olivares, Maricela Curisinche-Rojas, Margot Haydee Vidal-Anzardo, Johanny Fidela de Fátima Muro-Cieza, Karina Mayra Aliaga-Llerena de Nuñez.

## Data availability

The mortality data underlying the results of this study are available in the public domain from the WHO Mortality Database (https://www.who.int/data/data-collection-tools/who-mortality-database). The statistical analysis code and datasets are available from the corresponding author upon reasonable request.

## Data Availability

The mortality data underlying the results of this study are available in the public domain from the WHO Mortality Database (https://www.who.int/data/data-collection-tools/who-mortality-database). The statistical analysis code and additional supplementary files are available from the corresponding author upon reasonable request.

